# A Proposed Randomized, Double Blind, Placebo Controlled Study Evaluating Doxycycline for the Prevention of COVID-19 Infection and Disease In Healthcare Workers with Ongoing High Risk Exposure to COVID-19

**DOI:** 10.1101/2020.05.11.20098525

**Authors:** Paul A Yates, Ashton M Leone, Elias Reichel

**Affiliations:** Department of Ophthalmology, University of Virginia; Department of Ophthalmology, Tufts University

## Abstract

This paper proposes both a rationale and potential study design for evaluation of low dose doxycycline (20mg BID) for the prevention of COVID-19 infection in exposed health care workers. More generally, it provides a potential study design blueprint to other investigators for any interventional COVID-19 study looking to evaluate interventions for prevention or treatment of COVID-19 infection. This specific study described is a randomized, double blind, placebo controlled study to evaluate the efficacy and safety of doxycycline for the prevention of COVID-19 infection and disease in healthcare workers with ongoing high risk exposure to COVID-19. This study would consist of a 50-day Treatment Period (Day 0-Day 50), followed by an End of Study Visit, approximately 30 days after completion of study drug dosing. Initially, for approximately the first 4 to 6 weeks, an initial open-label arm would be enrolled with up to 1938 subjects who will be assigned to take 20mg doxycycline BID. In the double blind, placebo controlled arms approximately 3,692 participants would be randomized to either doxycycline or placebo for 50 days.

Doxycycline is a rational candidate drug to be evaluated for repurposing against SARS-CoV-2. Doxycycline is a generally safe tetracycline derivative that has been available for decades, most commonly dosed at 100mg BID to treat bacterial infections. However, in addition to its antimicrobial properties, doxycycline (and more generally tetracycline derivatives) may have a role as an effective anti-viral agent and as an anti-inflammatory drug. Early studies indicate potential efficacy of minocycline against respiratory syncytial virus (RSV) [12], and doxycycline against Dengue and Chikungunya infection[9, 10]. In addition, doxycycline is known or proposed to target several pathways that regulate viral replication. [13, 14, 15]. Doxycycline is a particularly attractive candidate as a COVID-19 prophylactic given it has been used in clinical practice for decades and maintains an excellent safety profile as demonstrated in multiple clinical studies. Any effective prophylaxis for COVID-19 should be able to demonstrate high efficacy at preventing infection and/or lowering severity of disease.

Equally important, it should demonstrate this efficacy at dosing levels that are highly unlikely to precipitate any untoward severe side effects. Doxycycline has been selected based on its ability to: 1) inhibit metalloproteinases (MMPs), implicated in initial viral entry into the cell as well as in acute respiratory distress syndrome (ARDS) associated with severe COVID-19 infection [13, 16]; 2) potential to inhibit Papain-like proteinase (PLpro) responsible for proteolytic cleavage of the replicase polyprotein to release non-structural proteins 1, 2 & 3 (Nsp1, Nsp2 and Nsp3) all essential for viral replication. [19]; 3) potential to inhibit 3C-like main protease (3CLpro) or Nsp5 which is cleaved from the polyproteins causes further cleavage of Nsp4-16 and mediates maturation of Nsps which is essential in the virus lifecycle. [19]; 4) act as an ionophore help transport Zinc intracellularly, increasing cellular concentrations of Zinc to inhibit viral replication. [6, 15]; 5) inhibit Nf-kB which may lower inflammatory response to COVID-19 infection, and lower risk of viral entry due to decreasing DPP4 cell surface receptor. [20, 21]; 6) inhibits (specifically low-dose doxycycline) expression of CD147/EMMPRIN that may be necessary for SARS-CoV-2 entry into T lymphocytes [22, 23].

## 1. Introduction

SARS-CoV-2 is a newly emerged coronavirus for which there is currently no available vaccine and no proven prophylactic treatment. Until a vaccine becomes available, there will be an ongoing need for prevention of infection through public health, clinical testing, behavioral, and pharmacological means. This is particularly true for health care workers who risk daily exposure to COVID-19 positive patients and community contacts. Unfortunately, personal protection equipment (PPE) has been shown insufficient in and of itself, with 2-6% of healthcare workers routinely exposed to COVID-19 patients becoming infected themselves [1, 2]

The quickest route to a therapy is to repurpose existing approved medications if they show some activity against COVID-19 to either block infection, or prevent viral replication as a means to lower the severity of disease of those who do become infected. In this regard, the repurposing of hydroxychloroquine and chloroquine for chemoprophylaxis has garnered the most attention. Early studies report hydroxychloroquine may actually substantially worsen outcomes in COVID-19+ patients, so it is difficult to recommend them as prophylactic therapy or treatment at this time [3]. National double blind, placebo controlled, well powered studies are currently underway to evaluate true efficacy of these medications. However, even if ultimately proven effective, there remains concern over the potential for significant cardiovascular and ocular toxic side effects with widespread use of these agents, and the NIH currently recommends against use of hydroxychloroquine therapy as a prophylactic [4, 5].

There are additional approved medications that have been proposed to have efficacy in preventing SARS-CoV-2 infection or lowering disease severity or duration in patients infected with COVID-19. In particular, doxycycline is a rational candidate drug to be evaluated for repurposing against SARS-CoV-2. Doxycycline is a generally safe tetracycline derivative that has been available for decades, most commonly dosed acutely at 100mg BID to treat bacterial infections. Lower dose, 20mg BID has been proven effective for longer term treatment (i.e. months or years) of acne rosacea and periodontal disease [6]. At the lower dose the primary observed side effects are short term nausea when first initiating treatment in a few subjects and sun sensitivity [7]. Our recent trial at the University of Virginia, involving 300 subjects (average age 80 years old) evaluating low dose doxycycline use over 2 years to prevent progression of macular degeneration, revealed no safety issues with the medication (TOGA study https://clinicaltrials.gov/ct2/show/NCT01782989). Beyond routine use as an anti-biotic, doxycycline (and more generally tetracycline derivatives) may have a role as effective anti-viral agents and as an anti-inflammatory drug that can ameliorate the inflammatory response following viral infection [8, 9, 10, 11]. Early studies indicate potential efficacy of minocycline against respiratory syncytial virus (RSV) [12], and doxycycline against Dengue and Chikungunya infection [9, 10]. In addition, doxycycline is known or proposed to target several pathways that regulate viral replication [13, 14, 15].

Potential mechanisms of action for doxycycline against COVID-19 include: 1) inhibition of metalloproteinases (MMPs), in particular MMP-9 which has been demonstrated important for viral replication [13, 16]; 2) inhibition of Nf-kb, which is known to inhibit both the cytokine inflammatory response such as IL-6 (aka cytokine storm), and also viral load in general [17, 18]; 3) proposed inhibition of papain-like proteinase (PLpro) responsible for proteolytic cleavage of the replicase polyprotein which releases non-structural proteins 1, 2 & 3 (Nsp1, Nsp2 and Nsp3) essential for viral replication [19]; 4) proposed inhibition of 3C-like main protease (3CLpro) or Nsp5 which is cleaved from the polyproteins, causing further cleavage of Nsp4-16, mediating maturation of Nsps essential in the virus lifecycle 30]; 5) inhibition of proper processing of replicase proteins and RNA dependent RNA polymerase (RdRp) activity. Doxycycline, like hydroxychloroquine, is an ionophore which binds divalent cations (including Zinc) to facilitate cell membrane transport, and thereby increase Zinc concentration in the cell. Elevated Zinc has a known inhibitory effect on the replication of SARS-CoV, and potentially many other viral types which is why it is frequently used for the common cold (some of which are caused by human coronavirus) [15]; 6) apoptosis of senescent epithelial cells, which have known increased expression of DPP4/CD26 which serves as an entry point for COVID-19 into the cell, and which likely exhibit increased viral replication in these cells [20]; 7) prevention of viral entry into the cell via inhibition of Nf-kb, which in turn binds to the DPP4 promoter thereby diminishing DPP4 cell surface expression required for viral entry [21]. Note that DPP4 demonstrates increased expression in older subjects and subjects with diabetes or pulmonary disease, and may account in part for their increased morbidity and mortality to COVID-19 as compared to younger/healthier demographics [20]; 8) inhibits (specifically low-dose doxycycline) expression of CD147/EMMPRIN which studies suggest is needed for SARS-CoV-2 entry into T lymphocytes [22, 23].

**Figure 1.**
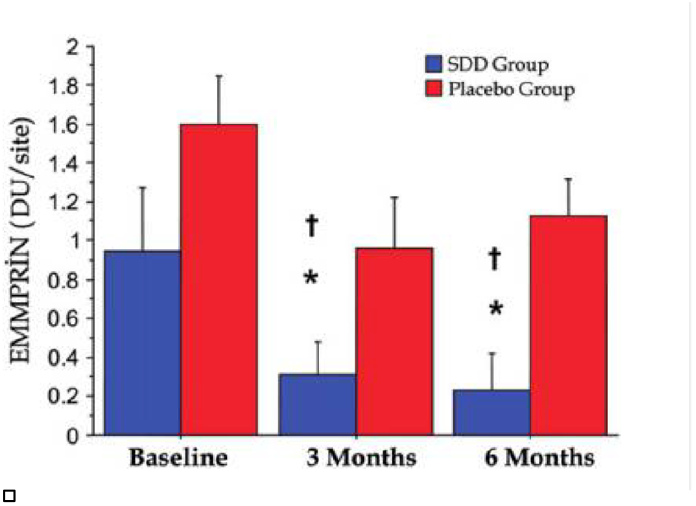
- sub-microbial doxycycline (SDD) substantially reduces CD147/EMMPRIN in subject gingival fluid. Expression of CD147/EMMPRIN on the cell surface may be required for viral entry of SARS-CoV-2

Doxycycline is particularly attractive amongst the many tetracycline derivatives, given that it has a clear dosing split between its cellular cytotoxic /anti-microbial effects (> 100 mg) and its anti-inflammatory/anti-viral/pro-ionophoric effects (40 mg) [6]. Thus, when administered twice a day on a large population basis it is highly likely to be well tolerated for the planned 50 day study Treatment Period, maintain its excellent safety profile as demonstrated in multiple clinical studies, while potentially inhibiting inflammatory and antiviral pathways important in initial infection, viral-replication, and ultimately severity and duration of disease for those that become infected. If proven efficacy is established in this clinical trial, the relatively benign side effects of low-dose doxycycline would render it amenable to population based treatments over the likely needed course of 18-24 months before a vaccine arrives. Benefit may be expected to far outweigh any potential risk.

The DOXYPRO trial will evaluate chemoprophylaxis with doxycycline for prevention of SARS-CoV-2 infection and decrease in severity and duration of disease of those subjects who become COVID-19 positive during the course of the study. Study subjects will be limited to healthcare workers with known COVID-19 negative status, with ongoing high risk exposure to COVID-19 positive patients. Potential study subjects meeting inclusion/exclusion criteria will be dosed at 20mg BID doxycycline, as compared to placebo BID in the control group. As many subjects are already taking dietary supplements such as Zinc, subjects already on a Zinc supplement will be allowed to remain on this supplement. Dosing will be 20mg doxycycline/placebo in morning and evening (dosed approximately 12 hours apart). If the subject is taking Zinc, we will advise them to take 50mg Zinc gluconate mid-day to prevent Zinc interference with systemic absorption of the doxycycline. This dosing regimen will eliminate chelation of Zinc by Doxycycline in the GI tract allowing time for absorption of both for those subjects on both. 3876 total COVID-19 negative subjects will be randomized in a 1:1 ratio to Doxycycline treatment (1938 subjects) and to placebo (1938 subjects). Subjects will remain on treatment or placebo for a total of 50 days to determine the safety and efficacy of low-dose, sub-microbial doxycycline to 1) reduce the infection rate of COVID-19 in previously uninfected subjects and 2) reduce the clinical severity of COVID-19 disease for those subjects that become infected during the course of the study. 3876 subjects are sufficient to power the study to have an 80% chance of detecting a statistically significant 2-fold difference in rate of infection of subjects on doxycycline versus placebo, assuming a 2.5% rate of infection in placebo subjects and 1.25% rate of infection of those taking doxycycline during the 50 day study.

A priori assumptions have to be made regarding normal expected infection rate in health care workers based on necessarily incomplete data. In support of our estimated incidence of infection, the Henry Ford Health care system recently reported approximately 700 workers, 2.3% of their health care work force of 30,000 people, tested COVID-19+ [1]. This number may of course be lower or higher depending on varying demographic, COVID-19+ case load and job function characteristics of enrolled health care workers. Given the urgent medical need, safety profile of doxycycline, as well as to provide better insight into recruitment and operational logistics for the study, we have elected to run a parallel initial open label arm in which subjects will be placed on 20mg low dose doxycycline. This open label study is anticipated to continue for approximately the first 4-6 weeks of enrollment, enrolling up to 1938 subjects, before the double blind, placebo controlled component is initiated upon completion of study drug/placebo manufacture and packaging. This arm will serve as a sentinel for any potential safety or efficacy signals in the study population and establish whether and to what degree the study has been appropriately powered. Similarly, many healthcare providers have self-initiated elemental Zinc, which may potentially, but not necessarily, affect study drug efficacy given that doxycycline serves as an ionophore. The open label arm may also provide further insight regarding any potential challenges or benefits to the concomitant use of Zinc supplements in addition to doxycycline, versus those not. To standardize treatment for this variable we will recommend those taking Zinc formulations use 50mg Zinc gluconate [24]. If they are not on that form of Zinc and prefer to stay on Zinc through the study, we will offer to supply the Zinc to reduce variability around the Zinc in the study. Results of this open-label doxycycline study will necessarily be analyzed separate from those of the double blinded, placebo controlled study.

### 1.1 Background

Among the tetracycline derivatives, doxycycline is an attractive candidate for safe prophylactic prevention of COVID-19 infection in exposed, high-risk subjects given a clear dosing split has been identified between its antimicrobial (>100 mg/day) and anti-inflammatory (40 mg/day) properties. Sub-antimicrobial doxycycline has found widespread clinical use for long-term suppression of acneiform and rosacea skin lesions with treatment effect dependent on its anti-inflammatory rather than anti-bacterial properties [25]. Sub-antimicrobial dose doxycycline has also found use in treating chronic periodontitis, proposed as a treatment for abdominal aneurysm, and as an adjunctive treatment in combination with methotrexate for rheumatoid arthritis [26-29]. Studies have shown no alterations in the composition of oral or intestinal microflora for subjects receiving long-term sub-antimicrobial doses of doxycycline [30-32].

### 1.2 Clinical Experience

Generic low-dose doxycycline (20mg, twice a day) is a tetracycline derivative most frequently prescribed for treatment of inflammatory lesions of rosacea in adults. ORACEA® is a branded, sustained release form of low-dose doxycycline (30mg immediate release, 10mg delayed release) also approved for rosacea, but dosed once per day [33, 34]. Rosacea is a chronic inflammatory disorder with characteristic skin lesions that include redness, visible blood vessels, papules and pustules that appear on the forehead, nose, and cheeks. The presumed mechanism of action is through reduction of skin inflammation rather than its antimicrobial properties, as 40mg doxycycline is known to be a sub-antimicrobial dose [35].

#### 1.2.1 Clinical Experience with Low Dose Doxycycline

ORACEA® is the best clinically studied formulation of low dose doxycycline, undergoing several phase III trials prior to FDA approval for treatment of rosacea. Del Rosso et al. [33, 34] compared ORACEA® versus placebo for treatment of rosacea in two (study 301 and 302) phase III, placebo controlled, double blind, randomized multicenter studies [10]. In Study 301, 32% of the subjects were 51 to 70 years of age and 33% in Study 302. In both studies, participants received ORACEA® (n = 269) or placebo (n = 268) once daily for 16 weeks. The mean number of inflammatory lesions at baseline was approximately 20 in both studies. By week 16, study 301 showed a decrease in inflammatory lesions of −11.8 in the treatment versus −9.5 in control groups. In comparison, study 302 showed a decrease of −5.9 compared to −4.3 in treatment versus control group (p< 0.001 for both studies).

In both study 301 and 302, ORACEA® was well-tolerated, with no major safety issues identified in the treatment group. For study 301, adverse events were reported in 44% of treatment versus 38.7% of control group participants, with most adverse events rated as mild or moderate in severity. Common adverse events included: diarrhea (4.8% treatment versus 3.3% control), nasopharyngitis (4.4% treatment versus 2.6% control), and headache (4.4% treatment versus 5.9% control). All subjects received hematology and serum chemistry panels at baseline and week 16 with no significant deviations or trends identified during the course of treatment.

A randomized, double blind, placebo controlled parallel group study evaluated the efficacy of adjunctive ORACEA® compared to placebo in adult subjects with untreated periodontitis. Subjects were treated by scaling and root planning (SRP) and were assigned to receive either ORACEA® or placebo once daily for 9 months. The study demonstrated ORACEA® as an adjunct to SRP achieved significantly greater clinical benefits compared to SRP alone [36].

Compliance with the once-daily dosing regimen was high (> 92%). ORACEA® was well tolerated, with no clinically meaningful difference in the numbers of adverse events reported by the treatment group compared to the placebo group, including adverse events associated with the genitourinary and gastrointestinal tracts or skin [36]. 88 subjects (66.2%) in the ORACEA® group reported 217 adverse events and the most common were: headache (9.8%) and influenza and nasopharyngitis (5.3% each). 94 subjects (70.7%) in the placebo group reported 229 adverse events and the most common were: sensitivity of teeth (9.8%) and headache and nasopharyngitis (7.5% each). No adverse event or serious adverse event in either group was evaluated by the investigators to be probably related to the study drug. There were no clinically meaningful differences between the treatment and placebo control groups with respect to laboratory tests (complete blood count and blood chemistry) or changes in laboratory test parameters between baseline and Month 9 [36].

With respect to microbiologic outcomes, the study demonstrates that longterm use of ORACEA® does not result in a change in microbial flora, an increase in doxycycline resistance, the acquisition of doxycycline resistance, or the emergence of cross-resistance or multi-antibiotic resistance [36].

#### 1.2.2 Long Term Clinical Experience with Sub-antimicrobial Doxycycline

Longer term interventions with sub-antimicrobial dose doxycycline have also been examined.

A double blind, placebo controlled trial that evaluated the intestinal flora in adult subjects with chronic periodontitis demonstrated long-term treatment with sub-antimicrobial dose doxycycline has no antibacterial effect on intestinal flora. 69 subjects (30 – 75 years of age) were randomized to receive either 20mg doxycycline (sub-antimicrobial dose) or placebo-control twice-daily for 9 months. Specifically, the results suggest a 9-month regimen of sub-antimicrobial dose doxycycline does not result in: a change in the normal fecal or vaginal flora, an increase in doxycycline resistance, the acquisition of doxycycline resistance, or the emergence of multiantibiotic resistance [37]

Four multicenter, placebo controlled, double blind, randomized clinical trials evaluated the administration of multiple doses of sub-antimicrobial dose doxycycline for the treatment of adult periodontitis. In studies 1-3, 437 subjects were randomized to receive placebo, 10 mg doxycycline daily, 20 mg doxycycline daily, or 20 mg doxycycline twice daily for 12 months. In study 4, 190 subjects were randomized to either placebo or 20 mg doxycycline twice daily for 9 months. A meta-analysis of all four studies showed doxycycline was well-tolerated and reported adverse events were similar between the doxycycline groups and placebo control group with no difference in the occurrence of adverse reactions usually associated with higher-dose tetracyclines between the groups [38]. No clinically significant differences were identified for liver or kidney function tests in the treatment groups as compared to the control group and no resistance developed to doxycycline during the course of the study [38].

A randomized, double blind clinical study compared the efficacy of doxycycline adjunctive to methotrexate (MTX) versus MTX alone in 66 adult subjects (27 – 74 years of age) with early seropositive rheumatoid arthritis (RA). Subjects were randomized to: 100 mg doxycycline (antimicrobial dose) twice-daily plus MTX, 20 mg doxycycline (sub-antimicrobial dose) twice-daily plus MTX, or placebo plus MTX for a period of two years. Both doxycycline treatment groups exhibited the same reduction in RA severity over the course of the study, and the improvement in both treatment groups was greater than the improvement exhibited by the placebo-control group [29]. Additionally, the number of adverse events reported in the sub-antimicrobial dose treatment group and the placebo group was equivalent and less than the number of adverse events reported by the antimicrobial dose treatment group [29].

A two year, double blind, randomized trial compared treatment with 20mg doxycycline (sub-antimicrobial dose) twice daily versus placebo in 128 post-menopausal women 45 – 70 years age (at the time of screening) with chronic periodontitis. Results showed a statistically significant reduction in serum inflammatory biomarkers hs-CRP and MMP-9 in the treatmentversus control group. Notably, there was no sign of microbiological resistance in the treatment group as compared to the placebo group and no significant safety issues were identified during the course of the study [39].

Finally, a two year, double blind, randomized trial compared treatment with ORACEA® versus placebo is ongoing in 286 subjects with advanced nonexudative age related macular degeneration (TOGA study https://clinicaltrials.gov/ct2/show/NCT01782989). The average subject age in this study is over 75. No significant safety issues have been identified in this aged population during the course of the study.

Clinical studies to-date with thousands of subjects using low-dose doxycycline have demonstrated efficacy across multiple indications as well as no significant safety issues. This clinical team also has direct experience with regards to the safety of doxycycline over the past 4 years as part of a clinical study in elderly subjects with macular degeneration (TOGA study https://clinicaltrials.gov/ct2/show/NCT01782989). Safety is extremely important for any proposed prophylactic treatment given the need to treat millions of subjects to reduce overall rates of infections. Efficacy of the proposed medication needs to be high, but equally important is having a robust safety profile for population based treatments. It is this specific requirement that has caused us to select low-dose doxycycline over antibiotic strength doxycycline for this study.

## 2. Study Objectives and Endpoints

### 2.1 Study Objectives

This study will assess the efficacy and safety of daily oral administration of Doxycycline Hyclate 20mg BID compared with daily oral administration of placebo in COVID-19 negative healthcare workers with known ongoing high risk exposure to COVID-19 positive patients to 1) prevent COVID-19 infection and 2) reduce severity of disease and duration of symptoms in those subjects who become infected with COVID-19.

#### 2.1.1 Primary Objective

- Evaluate the efficacy of daily oral administration of Doxycycline Hyclate 20mg BID as compared with placebo control over the course of 50 days to prevent COVID-19 infection in subjects with ongoing high risk exposure to COVID-19 positive patients.
- An infection by SARS-CoV-2 is defined by a positive specific Reverse Transcription - Polymerase Chain Reaction (RT-PCR) on nasopharyngeal swab.
  ○ At study initiation Potential subjects must have:
    ■ a documented negative Reverse Transcription - Polymerase Chain Reaction (RT-PCR) on nasopharyngeal swab
  ○ After study initiation, subjects will be deemed to have failed treatment if they demonstrate:
    ■ a positive specific Reverse Transcription - Polymerase Chain Reaction (RT-PCR) on nasopharyngeal swab during follow-up OR
    ■ a positive specific RT-PCR on a respiratory sample in case of onset of symptoms consistent with COVID-19 during follow-up OR
    ■ a seroconversion to SARS-CoV-2 after randomization.

#### 2.1.2 Secondary Objectives

- Evaluation of the incidence of symptomatic SARS-CoV-2 infections will be completed in all study arms. Symptomatic is defined as positive RT-PCR on nasopharyngeal swab with clinical symptoms consistent with presumed COVID-19 infection.
  - Clinical symptoms consistent with presumed COVID-19 infection are defined as:
    - fever
    - tiredness
    - dry cough
    - anosmia
    - aches and pains
    - nasal congestion
    - rhinorrhea
    - sore throat
    - diarrhea.
    - dyspnea
    - conjunctival congestion
    - sputum production
    - headache
    - hemoptysis
    - vomiting
- Evaluation of the incidence of asymptomatic SARS-CoV-2 infections in treatment and placebo group. Asymptomatic is definedas positive RT-PCR from nasopharyngeal swab without any clinical symptoms associated with COVID-19 infection.
- Evaluation of the incidence of mild, moderate, severe, and critical cases of SARS-CoV-2 infection in all study arms.
  - Mild, moderate, severe and critical SARS-CoV-2 infections among health-care workers in each arm are defined by:
    - Mild to moderate:
      - a positive RT-PCR for COVID-19 infection from nasopharyngeal swab
      - Clinical symptoms of SARS-CoV-2 infection not progressing to Severe
    - Severe
      - a positive specific RT-PCR on a respiratory sample OR
      - a thoracic CT scan with imaging abnormalities consistent with COVID-19 performed in case of onset of symptoms consistent with COVID-19 during follow-up in a participant who need to be hospitalized for respiratory distress. Respiratory distress defined as dyspnea with a respiratory frequency > 30/min, blood oxygen saturation <93%, partial pressure of arterial oxygen to fraction of inspired oxygen ratio <300 and/or lung infiltrates >50%
    - Critical:
      - respiratory failure as defined for severe requiring ventilation
      - septic shock
      - multiple organ dysfunction/failure
      - death
- Evaluate the safety of daily oral administration of the investigational drug through the collection of adverse events and serious adverse events, vital signs measurement, and clinical laboratory assessments.
- Evaluation of the study completion rate of the study in both treatment and placebo arms.
- Evaluation of the discontinuation rate of the investigational drug in the treatment arm.
- Evaluation of the adherence of study participants to study drug and dosing regimen by self-assessment and count of study drug. This will include subjects who test COVID-19+ during the duration of the study as they will be asked to continue to take study drug through Day 50 if able and medically advisable per their care provider.
- Evaluation of the possible effects of Zinc supplementation on the prevention of COVID-19 infection and reduction of severity of disease and duration of symptoms in those subjects who become infected with COVID-19.

## 3. Study Endpoints

### 3.1.1 Primary Endpoint

1. Number of enrolled subjects who convert from COVID-19 negative to COVID-19 positive status during the course of the 50 day Treatment Period in each study arm. Outcome metric will be expressed as a relative risk ratio for COVID-19 infection of subjects taking blinded doxycycline to subjects taking blinded placebo.

### 3.1.2 Secondary Endpoints

1. Incidence of symptomatic SARS-CoV-2 infections in each study arm during Treatment Period. Symptomatic is defined as positive RT-PCR on nasopharyngeal swab with clinical symptoms consistent with presumed COVID-19 infection.
2. Incidence of asymptomatic SARS-CoV-2 infections in each study arm during Treatment Period. Asymptomatic is defined as positive RT-PCR from nasopharyngeal swab without any clinical symptoms associated with COVID-19 infection.
3. Incidence of symptomatic SARS-CoV-2 infections in each study arm in 30 days following Treatment Period. Symptomatic is defined as positive RT-PCR on nasopharyngeal swab with clinical symptoms consistent with presumed COVID-19 infection.
4. Incidence of asymptomatic SARS-CoV-2 infections in each study arm in 30 days following Treatment Period. Asymptomatic is defined as positive RT-PCR from nasopharyngeal swab without any clinical symptoms associated with COVID-19 infection.
5. Evaluation of the incidence of mild to moderate, severe, and critical SARS-CoV-2 infections in each study arm.
6. Time-to-first SARS-CoV-2 infection as defined by positive RT-PCR from nasopharyngeal swab in each study arm.
7. Time-to-first SARS-CoV-2 clinical event consisting of a persistent change for any of the following: (1) positive nasopharyngeal swab (2) clinical symptoms of COVID-19 infection in each study arm
8. Time-to-first SARS-CoV-2 clinical worsening from asymptomatic to mild to moderate in each study arm.
9. Time-to-first SARS-CoV-2 clinical worsening from mild to moderate to severe in each study arm.
10. Rate of subject reported adverse events in each study arm.
11. Percentage of subjects who complete the 50 day Treatment Period in each study arm.
12. Percentage subjective compliance of subjects with specified study drug administration schedule in each study arm.
13. Percentage objective compliance (as measured by pill counts) of subjects with specified study drug administration schedule in each study arm.

### 3.1.3 Safety Endpoints

1. Incidence and severity of systemic adverse and serious adverse events in all treatment arms.
2. Changes and abnormalities in clinical laboratory parameters in all treatment arms.
3. Changes in vital signs in all treatment arms.

### 3.1.4 Subset Analysis

For all subjects, subset analysis will be performed for those subjects taking a Zinc supplement as compared to those not on Zinc supplementation.

## 4. Study Design

### 4.1 Overview

This is a randomized, double blind, placebo controlled study to evaluate the efficacy and safety of doxycycline for the prevention of COVID-19 infection and disease in healthcare workers with ongoing high risk exposure to COVID-19. This study consists of a 50-day Treatment Period (Day 0-Day 50), followed by an End of Study Visit, approximately 30 days after completion of study drug dosing. Initially, for approximately the first 4 to 6 weeks, an initial open-label arm will be enrolled with up to 1938 subjects who will be assigned to take 20mg doxycycline BID. In the double blind, placebo controlled arms approximately 3,692 participants will be randomized to either doxycycline or placebo for 50 days.

### 4.2 Rationale for Study

Doxycycline is a rational candidate drug to be evaluated for repurposing against SARS-CoV-2. Doxycycline is a generally safe tetracycline derivative that has been available for decades, most commonly dosed at 100mg BID to treat bacterial infections. However, in addition to its anti-microbial properties, doxycycline (and more generally tetracycline derivatives) may have a role as an effective anti-viral agent and as an anti-inflammatory drug. Early studies indicate potential efficacy of minocycline against respiratory syncytial virus (RSV) [12], and doxycycline against Dengue and Chikungunya infection[9, 10]. In addition, doxycycline is known or proposed to target several pathways that regulate viral replication. [13, 14, 15]

Doxycycline is a particularly attractive candidate as a COVID-19 prophylactic given it has been used in clinical practice for decades and maintains an excellent safety profile as demonstrated in multiple clinical studies. Any effective prophylaxis for COVID-19 should be able to demonstrate high efficacy at preventing infection and/or lowering severity of disease. Equally important, it should demonstrate this efficacy at dosing levels that are highly unlikely to precipitate any untoward severe side effects.

Doxycycline has been selected based on its ability to: 1) inhibit metalloproteinases (MMPs), implicated in initial viral entry into the cell as well as in acute respiratory distress syndrome (ARDS) associated with severe COVID-19 infection [13, 16]; 2) potential to inhibit Papain-like proteinase (PLpro) responsible for proteolytic cleavage of the replicase polyprotein to release non-structural proteins 1, 2 & 3 (Nsp1, Nsp2 and Nsp3) all essential for viral replication. [19]; 3) potential to inhibit 3C-like main protease (3CLpro) or Nsp5 which is cleaved from the polyproteins causes further cleavage of Nsp4-16 and mediates maturation of Nsps which is essential in the virus lifecycle. [19]; 4) act as an ionophore help transport Zinc intracellularly, increasing cellular concentrations of Zinc to inhibit viral replication. [6, 15]; 5) inhibit Nf-kB which may lower inflammatory response to COVID-19 infection, and lower risk of viral entry due to decreasing DPP4 cell surface receptor. [20, 21]; 6) inhibits (specifically low-dose doxycycline) expression of CD147/EMMPRIN that may be necessary for SARS-CoV-2 entry into T lymphocytes [22, 23].

### 4.3 Rationale for Dosage

20mg BID dosing is chosen because this dose of doxycycline is sub-antimicrobial, will not induce antibiotic resistance, and has known high subject safety while maintaining the anti-inflammatory, and potentially anti-viral ionophoric capabilities of the drug. The study requires twice-daily administration which is consistent with the prescribing recommendations for sub-antimicrobial doxycycline for treatment of acne rosacea and periodontal disease. Additionally, prior studies indicate that long-term daily administration of sub-antimicrobial dose doxycycline is well tolerated in older subject populations [29, 34, 37, 39]

### 4.4 Description of Study Design

#### Open-label arm

The open-label arm will be enrolled during approximately the first 4-6 weeks of the study. After providing informed consent, potential subjects will undergo screening (Day 0) to determine if they are eligible to participate in the study. If eligible, a CoViD-19 – test will be required following screening and prior to starting study drug. After eligibility is confirmed, eligible subjects will be assigned to known sub-antimicrobial doxycycline treatment group. Up to 1938 eligible subjects will be enrolled at Day 0 to complete the study Treatment Period and End-of-Study Follow-up Visit.

During the 50-day Treatment Period (Day 0 – Day 50), Site staff will be asked to contact the subjects via internet telemedicine visit at Day 28 (±3 days) and Day 50 (±3 days). 30 days (± 7 days) after completion of the Day 50 visit, participants will be asked to complete an End-of-Study Follow-up Telephone Visit. Subjects who have not become symptomatic and tested COVID-19+ during the Treatment Period, will take a COVID-19 test at the end of the Treatment Period so that we might detect subjects who are asymptomatic but remain COVID-19+. If a reliable (high sensitivity, high specificity) antibody test becomes available, we will, where available, confirm infection status with this test.

Subjects who discontinue prematurely from the study drug treatment will be encouraged to continue to comply with the study follow-up and visit schedule. Subjects who initiate study drug treatment and discontinue early from the study will be asked to complete an Early Termination Telephone Visit.

#### Placebo controlled, double blind arms

The placebo controlled, double blind study will begin enrollment 4 to 6 weeks after the open-label study commences. After providing informed consent, potential subjects will undergo screening (Day 0) to determine if they are eligible to participate in the study. If eligible, a COVID-19 – test will be required following screening and prior to starting study drug. After eligibility is confirmed, eligible subjects will be randomized in a 1:1 ratio to either the sub-antimicrobial doxycycline treatment group or placebo treatment group. Approximately 3876 eligible subjects (1938 subjects doxycycline/ 1938 subjects placebo treatment group) are randomized at Day 0 to complete the study Treatment Period and End-of-Study Follow-up Visit.

During the 50-day Treatment Period (Day 0 – Day 50), Site staff will be asked to contact the subjects via internet telemedicine visit at Day 28 (±3 days) and Day 50 (±3 days). 30 days (± 7 days) after completion of the Day 50 visit, participants will be asked to complete an End-of-Study Follow-up Telephone Visit. Subjects who have not become symptomatic and tested COVID-19+ during the Treatment Period, will take a COVID-19 test at the end of the Treatment Period so that we might detect subjects who are asymptomatic but remain COVID-19+. If a reliable (high sensitivity, high specificity) antibody test becomes available, we will, where available, confirm infection status with this test.

Subjects who discontinue prematurely from the study drug treatment will be encouraged to continue to comply with the study follow-up and visit schedule. Subjects who initiate study drug treatment and discontinue early from the study will be asked to complete an Early Termination Telephone Visit.

### 4.5 Flowchart of Randomized, Placebo controlled, Double blind Study Design

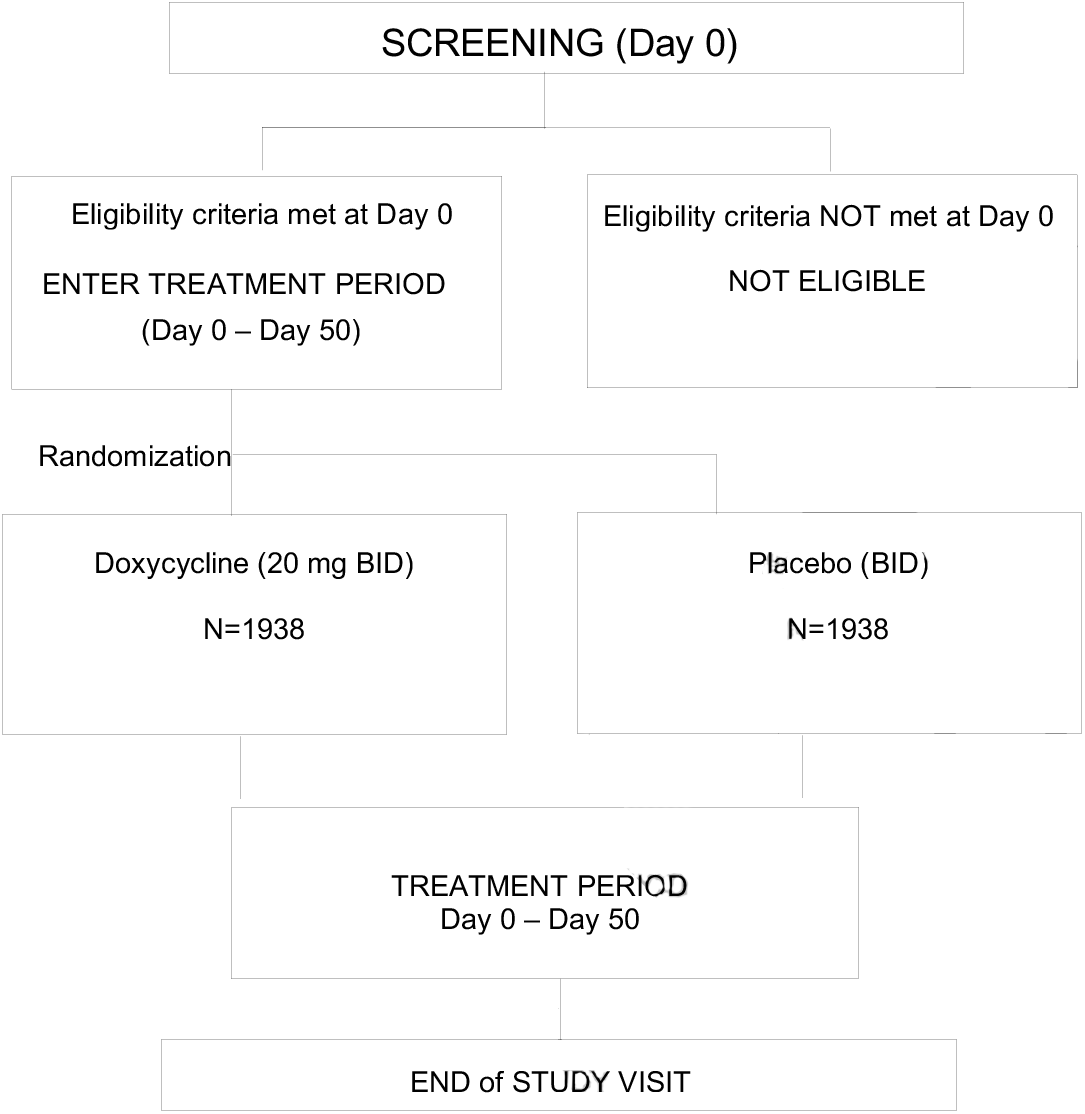

## 5. Study Population

### 5.1 Inclusion Criteria

#### General

1. Male or female, age 18 years or older.
2. Ability to swallow oral medications.
3. If a female of childbearing potential, must agree to use an effective form of contraception for the duration of the study.
4. Negative pregnancy test during the previous 7 days to start treatments or more than 2 years after menopause.
5. Willing and able to sign the informed consent.
6. Willing and able to comply with all required study visits and assessments/procedures.
7. Access to device and internet for Telehealth visits
8. Willing and able to receive study drug by mail
9. Willing and able to return the internet-based study questionnaires at Day 0, Day 50, and Day 80

#### COVID-19 Specific

10. High risk healthcare provider defined as someone who is actively engaged in working directly with and has direct contact with COVID-19 patients.
11. Must have a COVID-19 negative test after consent and screening are complete.

### 5.2 Exclusion Criteria

#### General

1. Pregnancy or women who are breast feeding
2. Subjects that lack decision-making capacity

#### Tetracycline-Derivative Specific Conditions

3. History of any hypersensitivity to tetracycline components.
4. Use of a tetracycline derivative for concurrent systemic disorder
5. Concurrent use of anti-coagulants, penicillin, methoxyflurane, oral retinoids, barbiturates, carbamazepine, and phenytoin.
6. History of sensitivity to the sun.
7. Use of any investigational or non-registered drug or vaccine within 30 days preceding the first dose of the study drugs, or planned use during the study period

#### Systemic Conditions

8. Concurrent or ongoing treatment for any systemic infection within 14 days of Day 0.
9. History of myocardial infarction, cerebrovascular accident, transient ischemic attack within 90 days of Day 0.
10. Known chronic kidney disease, stage 4 or 5 or receiving dialysis.
11. Known immunosuppressive condition or hematological disease
12. Symptomatic with subjective fever, cough, or sore throat
13. Current hospitalization

### 5.2 Screening / Day 0 Visit

The following exams and tests will be completed at the Screening / Day 0 Visit:

- Obtain Video Informed Consent over HIPAA compliant WebEx
- Demographics
- Medical and Surgical History Review
- Concomitant Medication Review
- COVID-19 Nasopharyngeal Viral Test
- Review Study Inclusion and Exclusion Criteria
- Determination of Participant Eligibility

Screening / Day 0 assessments may be completed over 24 hours but must be completed within 7 days of signing the informed consent. If a subject completes the nasopharyngeal viral test per standard practice prior to the signing of the informed consent, that test may be used to determine the subject’s eligibility if occurring within 24 hours of signing the informed consent. A negative serology for SARS-CoV-2 does not substitute for a nasopharyngeal viral test given the current variability in reported accuracy of these tests.

A subject must meet all the eligibility criteria at Screening / Day 0 to be considered eligible for randomization in the study.

#### 5.2.1 Randomization

All subjects enrolling in the double blind, placebo controlled phase of the clinical trial will be randomized after confirmation of eligibility from the Screening / Day 0 evaluation. Subjects will be randomized in a 1:1 ratio to doxycycline tablets or matching placebo tablets, to be taken orally twice per day for 50 days.

##### Randomization Details

The randomization list will be generated at the by the Department of Public Health Sciences, Division of Biostatistics. A combined total of 3876 treatment assignments will be generated. Subjects will be stratified by several variables including age, sex, race, geographic location, severity of reported cases in that geographic location at the time of enrollment and use of supplemental Zinc. The randomization list for each will be generated via a permuted-block randomization scheme that will require the treatment assignments to be in approximate balance at any point in time during the enrollment period. To avoid assignment discovery, the permuted-block size utilized in generating the randomization list will be random. Each permuted-block of assignments will include an equal number of doxycycline and matching placebo tablet assignments and the assignments will be sequentially utilized according to the permuted assignment order. The Division of Biostatistics will send the blind-protected (i.e. coded) randomization list to the Coordinating Center, where the lists will be maintained. Blind-protected coded drug bottles and matched coded treatment assignments will be used to provide study drug for each subject. Overnight courier will be used to deliver the coded drug bottle to each participant. Following study completion, and only after the Coordinating Center central database has been officially locked in preparation for data analysis, will the Division of Biostatistics reveal the treatment assignment code to the investigators and subjects. A backup, password protected copy of the original randomization list will be stored on the Information Technology and Communication Multi-Tier server. Two Biostatisticians from the Department of Public Health Sciences, Division of Biostatistics will have access to the password.

#### 5.2.2 Study Drug Distribution

Following confirmation of eligibility, the subject will be given their study drug with instructions for use. The study will utilize 20 mg doxycycline tablets or matching placebo tablets. Subjects should take one tablet each morning on an empty stomach (defined as at least one hour before or two hours after a meal) and one tablet each evening on an empty stomach. Subjects will be asked to start taking study drug the morning after they receive it by mail, and this will be established as Day 0. If a subject feels symptomatic between Day 0 of randomization and their starting the study drug they should obtain a confirmatory COVID 19 test. Administration of adequate amounts of fluid along with the tablets is recommended to wash down the tablet to reduce the risk of esophageal irritation and ulceration. The subject will continue with the twice-daily dosing regimen of the doxycycline for the duration of the 50 day Treatment Period.

Any subjects taking Zinc as part of their current pre-existing medication regimen will be directed to do so at mid-day, approximately four hours after the first doxycycline or matching placebo tablet. This is necessary to avoid unnecessary chelation of the study medication and Zinc, which would result in decreased systemic absorption for both. Any subject that is on Zinc supplements will be directed to take only 1 tablet of 50mg Zinc gluconate, once a day at mid-day, rather than some alternate Zinc formulation. If they cannot source 50mg Zinc gluconate, this will be provided to them along with study drug to maintain consistency in dose and formulation. If a subject desires to take a Zinc supplement, it must be for the duration of the study from Day 0 to Day 50.

### 5.3 Follow-up Telephone Calls and Telemedicine Visit: Day 28 and Day 50 (± 3 days)

The site will contact the subject to review potential adverse events, concomitant medications, and study drug compliance. The subject will obtain a COVID-19 test at Day 50 (± 3 days). If an antibody test is feasible we will also request the subject obtain an antibody test for SARS-CoV-2 at Day 50 (± 3 days).

### 5.4 End-of-Study Visit (30 Days ± 7 days after completion of Day 50 Visit)

The following exams and tests will be completed at the End-of-Study visit:

- Concomitant Medication Review
- Adverse Event Review

The End-of-Study Visit concludes the subject’s participation in the study.

### 5.5 Unscheduled Visits

An unscheduled telemedicine visit may be performed at any time during the study at the subject’s request or the request of the Medical Monitor. Exams and tests to be performed at the visit will be determined by the investigator and are dependent upon the nature of the visit. At a minimum, the following assessments are required at each Unscheduled Visit:

- Concomitant Medication Review
- Adverse Event Review
- Study Drug Intake Review

Subjects who become symptomatic with possible COVID-19 symptoms during the Treatment Period, should perform a COVID-19 test and adhere to the policies of their local institution and state.

### 5.6 Early Termination Visit

A subject may withdraw from the study at any time. The principal investigator, medical monitor or sponsor may also withdraw the subject from the study in the event of an intercurrent illness, adverse event, or other reasons concerning the health or well-being of the subject. The subject may also be withdrawn due to noncompliance with the protocol, a protocol violation, or other administrative reasons.

Subjects who are withdrawn prior to initiating the study drug, do not need to complete an Early Termination Visit. Subjects who are withdrawn after initiating the drug should complete an Early Termination Visit. All tests and procedures required at the End of Study Visit should be completed at the Early Termination Visit. These tests and procedures include:

- Concomitant Medication Review
- Adverse Event Review
- Study Drug Intake Review

Early Termination Visit (if conducted in place of the End-of-Study Visit) concludes the subject’s participation in the study.

### 5.7 Permanent Discontinuation of the Study Drug with Continuation in the Study

Subjects may permanently discontinue the study drug (due to subject request or investigator or sponsor discretion) due to intercurrent illness, adverse event, or other reasons concerning the health or well-being of the subject. Subjects who permanently discontinue the study drug should be encouraged to remain in the study (but off the study drug) and to complete all remaining follow-up visits and assessments.

Permanent discontinuation of the study drug is discussed further in Section 6.3.7.

## 6. Safety Considerations

A summary of known adverse drug reactions and drug interactions associated with doxycycline and tetracycline derivatives follows as well as plans to mitigate these events during the course of the study. Full prescribing information for doxycycline is included as Appendix C of this protocol.

### 6.1 Adverse Drug Reactions

In clinical trials of adult subjects with periodontal disease 213 subjects received 20mg BID over a 9 - 12 month period [40]. The most frequent adverse reactions occurring in studies involving treatment with doxycycline hyclate tablets or placebo are listed below:

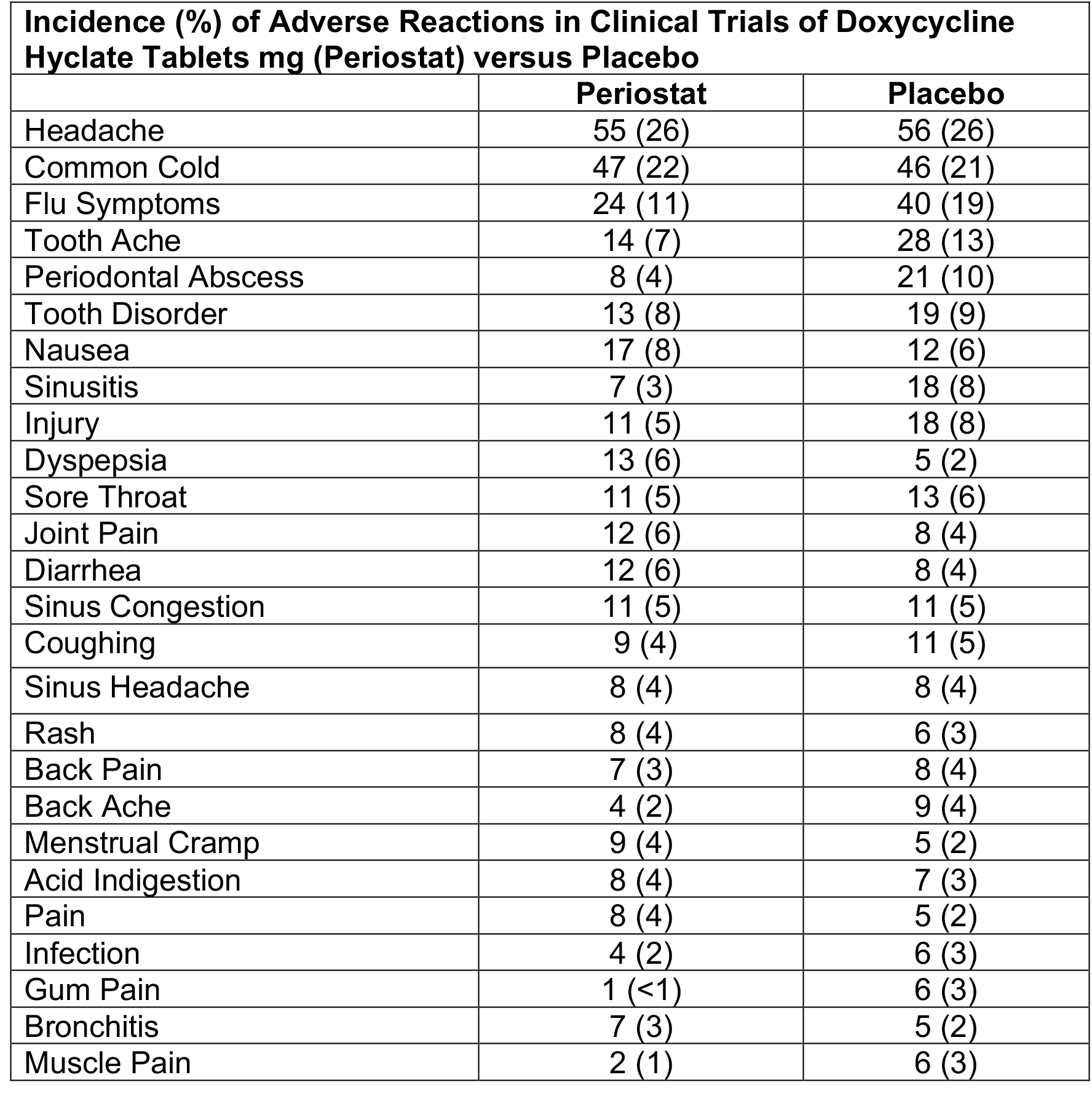

The following adverse reactions have been observed in subjects receiving tetracycline derivatives at higher, anti-microbial doses [7]. Because these events were reported in subjects receiving higher doses than required by this study, these events are unlikely, but should be noted:

#### Gastrointestinal

Anorexia, nausea, vomiting, diarrhea, glossitis, dysphagia, enterocolitis, and inflammatory lesions (within vaginal candidiasis) in the anogenital region. Hepatotoxicity is rare. Esophagitis and esophageal ulcerations are rare and have been reported in subjects receiving the tablet forms and occurred when the subject took their medication immediately prior to laying down.

#### Skin

Maculopapular and erythematous rashes and in rare instances exfoliative dermatitis have been reported. Photosensitivity has been observed in individuals taking tetracycline derivatives, but was not observed in participants in clinical studies evaluating low dose doxycycline.

#### Renal toxicity

An increase in BUN has been reported and appears to be dose-related.

#### Hypersensitivity reactions

Urticarial, angioneurotic edema, anaphylaxis, anaphylactoid purpura, serum sickness, pericarditis, and exacerbation of systemic lupus erythematosus have been reported.

#### Blood

Hemolytic anemia, thrombocytopenia, neutropenia, and eosinophilia have been reported.

#### Other

Tetracyclines have been associated with the development of pseudomembranous colitis, tissue hyperpigmentation, autoimmune syndromes, pseudotumor cerebri, drug-resistant bacteria, superinfection (although not observed in clinical studies evaluating low dose doxycycline), and fetal harm when administered to pregnant women [37].

### 6.2 Drug Interactions

Adverse reactions have been observed during the concurrent use of tetracycline derivatives and anticoagulants, penicillin, methoxyflurane, antacids and iron preparations, low dose oral contraceptives, oral retinoids, and barbiturates and anti-epileptics [7]. Specific recommendations with respect to concurrent use of these medications by study subjects are outlined in Section 7.

### 6.3 Safety Plan

#### 6.3.1 Querying for Adverse Events

Adverse event querying will occur at all study visits (after initiation of study drug), including unscheduled study visits. Additional tests and examinations may be required depending on the adverse events reported by the subject. All adverse events, regardless of the relationship to the study drug, will be recorded.

#### 6.3.2 Review of Concomitant Medications

A review of concomitant medications will occur at all study visits, including unscheduled study visits. At all study visits, subjects should be reminded of prohibited concomitant medications. Prohibited concomitant medications are discussed in Section 7.0.

#### 6.3.3 Assessment of Study Drug Compliance

Upon completion of the study or following Early Termination the Investigator or designated site staff will assess the subject’s compliance with study drug intake.

#### 6.3.4 Permanent Discontinuation of the Study Drug with Continuation in the Study

Subjects may permanently discontinue the study drug (due to subject request or investigator or sponsor discretion) due to intercurrent illness, adverse event, or other reasons concerning the health or well-being of the subject. The investigator should discuss the case with the Medical Monitor prior to making the decision to discontinue the study drug.

The study drug should be permanently discontinued immediately if there is evidence of any of the following:

- Pregnancy
- Pseudomembranous colitis
- Severe Photosensitivity
- Development of autoimmune disorde
- Renal or liver toxicity (≥ 3.0× ULN)
- Allergic reaction to the study drug

Subjects who permanently discontinue the study drug should be encouraged to remain in the study (but off the study drug) and to complete all remaining follow-up visits and assessments.

#### 6.3.5 Temporary Discontinuation of the Study Drug with Continuation in the Study

Temporary discontinuation of the study drug may be allowed in the event of an adverse reaction in which the Investigator identifies a possible causal relationship to the study drug and further evaluation of the adverse reaction is required. The study drug may be permanently discontinued or re-started pending the results of the additional evaluation(s) to determine if there is a causal relationship. All cases involving a potential temporary discontinuation or re-starting of study drug should be discussed with the Medical Monitor.

## 7. Concomitant Medications

All concomitant medications should be used in accordance with approved labeling and as prescribed throughout the duration of the study.

Adverse reactions have been observed when tetracycline derivatives are used concurrently with certain medications. The following medications should not be used by subjects during their participation in the study:

- Anticoagulants, with the exception of ≤ 325mg aspirin
- Penicillins
- Methoxyflurane
- Oral retinoids, including isotretinoin and acitretin
- Barbiturates, carbamazepine, and phenytoin

Other than the study drug, concurrent use of any systemic tetracycline derivative (e.g. tetracycline, doxycycline, minocycline) at any dose is not allowed for the duration of the study.

Doxycycline can cause oral contraceptives to be less effective. Low dose oral contraceptives should not be used during the study unless with a second form of contraception.

Proton pump inhibitors, antacids daily vitamins, or nutritional supplements containing aluminum, calcium, Zinc, or magnesium and iron-containing preparations may impair doxycycline absorption and should be taken at least 2 hours prior to or 3 hours after taking the study medication.

## 8. Concurrent Medical Conditions

All concurrent medical conditions (whether new or worsening) and present prior to or at Day 0 (prior to initiating study drug) must be recorded in the Case Report Forms (CRFs) as concurrent medical conditions. All medical conditions, whether new or worsening and present after the initiation of the study drug, must be recorded in the CRFs as adverse events, regardless of severity or assessed relationship to the study drug, and reported to the UVA Institutional Review Board (IRB) according to IRB guidelines. Collection of adverse events ends once the subject completes the End-of-Study Visit or Early Termination Visit (whichever occurs first). Refer to Section 14 for a description of adverse events and adverse event reporting requirements.

In the event the subject experiences a concurrent medical condition or adverse event (either new or worsening) during participation in the study, the subject may continue in the study with study drug treatment, unless the investigator or Medical Monitor determines the subject should cease taking the study drug and/or be withdrawn from the study. A decision to withdraw a subject from the study should only be made after consulting with the Medical Monitor (as described in Section 12).

The clinical course of management of the concurrent medical condition should be recorded on the appropriate Case Report Forms (CRFs).

## 9. Study Drug Administration

### 9.1 Study Drug

The study drug, 20mg doxycycline tablets are FDA approved, hard white tablets containing doxycycline with binding elements. We will supply the active drug and matching placebo comparator. The study drug bottles and drug kits will be packaged and labeled according to the randomization scheme. Packaged drug will be shipped to the Coordinating Center who will ship the drug to the study participants.

The study drug will be dispensed to the subject with instructions for use. Subjects should take on an empty stomach (defined as at least one hour before or two hours after a meal) one (1) tablet each morning and one (1) tablet each evening. Administration of adequate amounts of fluid along with the tablets is recommended to wash down the tablet to reduce the risk of esophageal irritation and ulceration. The subject will continue with the twice-daily (2 tablet) dosing for the duration of the 50-day Treatment Period, with the first dose occurring the morning after study drug is received.

If the subject is taking a Zinc supplement, the subject should take one (1) tablet of 50mg Zinc gluconate at noon, at least 4 hours removed from administration of the study drug. Only subjects that are currently on a Zinc supplement at the time of enrollment should continue on this if they desire to do so and taking of this supplement should continue for the duration of the 50-day Treatment Period. If the subject is on a Zinc supplement other than 50mg Zinc gluconate, they will be asked to switch to this formulation and dose on this schedule for the duration of the 50-day Treatment Period. If they are unable to source the 50mg Zinc gluconate, they will be requested to either a) discontinue their use of supplemental Zinc or b) we will provide the 50mg Zinc gluconate to them. Stratification, randomization and end of study analysis of subjects will account for the possible treatment effect of subjects taking versus not taking Zinc supplements during the Treatment Period.

At all study visits (scheduled and unscheduled) occurring after the initial distribution of the study drug, the Investigator or designated personnel should assess the subject’s compliance with the study drug regimen (this will be done via subject self-report and tablet counting and recording).

### 9.2 Study Drug Storage

Subjects should be instructed to store the study drug in a secure and safe area that is out of reach to children. Subjects should be instructed that study drug should not be shared with anyone, including other study subjects. The subject should store the study drug at room temperature 59°F to 86°F (15°C to 30°C), in a tightly enclosed container (the study drug bottle is acceptable) and out of light.

### 9.3 Study Drug Accountability

The Investigator or designated personnel must maintain accurate and complete records (including dates) of receipt of all study drug supplies. Drug accountability Case Report Forms must be completed by the Investigator or designated personnel to document the dispensing of the study drug to subjects.

### 9.4 Randomization and Study Drug Assignment

Study drug kits will be pre-labeled with a kit number corresponding to the randomization plan generated by the Department of Public Health Sciences, Division of Biostatistics. Each subject drug bottle will be assigned its own unique bottle number and individually labeled to correspond with the unique bottle number.

The treatment assignments will be randomly assigned to the subjects. A permuted-block randomization scheme will be used to ensure approximate balance between treatment assignments (see Section 5.2.1 for details). Once a subject has been assigned a bottle number, that bottle cannot be assigned to another subject.

### 9.5 Study Drug Masking

For the double blind, placebo controlled phase of the proposed clinical trial, all subjects, all study personnel, and the Coordinating Center will remain blind to the study drug treatment assignments until the conclusion of the entire study (see Section 5.2.1 for details). Only the designated Biostatisticians (from the Department of Public Health Sciences; Division of Biostatistics) will have access to the treatment assignments, but may not communicate the treatment assignments to any other study personnel at the Coordinating Center, until conclusion of the entire study, and the central database has been officially locked.

In the rare event an emergency unmasking of a subject treatment assignment is required, the procedures outlined in Section 10 will be followed.

## 10. Emergency Unmasking of Subject Treatment Assignment

Emergency unmasking of an individual subject’s treatment assignment is only allowed in the rare case of a medical emergency where the Investigator (or Medical Monitor) believes the treatment assignment must be revealed to ensure subject safety.

In this situation, the Investigator should contact the study Medical Monitor to make a request to unmask the subject. The Medical Monitor will make the final decision as to if emergency unmasking is allowed. If the Medical Monitor approves the Investigator’s request to unmask the subject then the Investigator may proceed with unmasking. If another situation arises in which the Investigator believes emergency unmasking is warranted, the situation must be discussed with the Medical Monitor and approval from the Medical Monitor is required prior to unmasking.

The subject’s treatment assignment will be revealed by one of the two Biostatisticians from the Department of Public Health Sciences, Division of Biostatistics who have access to the randomization list. The treatment assignment should only be shared with individuals involved in the direct care of the subject who may require this information to make medical decisions and should not be shared with the Coordinating Center.

If the emergency unmasking of an individual subject’s treatment assignment occurs, the Investigator must document the purpose, date, and personnel involved in the unmasking (and communication of the treatment assignment information).

## 11. Subject Withdrawals and Subjects Lost to Follow-up

In the event a subject self-elects to discontinue the study drug or the Investigator (after consulting the Medical Monitor) elects to discontinue the study drug, every effort should be made by the Investigator and / or designated site personnel to encourage the subject to continue to complete all protocol required study visits and assessments and not withdraw from the study. However, a subject may withdraw from the study at any time.

### 11.1 Subject Withdrawals

All subjects will be advised via the written informed consent form and informed consent process that they have the right to withdraw from the study at any time or may be withdrawn at the discretion of the Investigator, Medical Monitor, or Coordinating Center. Once a subject terminates the study prematurely (either by subject decision or Investigator, Medical Monitor, or Coordinating Center discretion), every effort should be made to complete an Early Termination Visit to evaluate the subject’s clinical status. For all subjects who terminate the study early, a reason for the subject’s early discontinuation should be documented on the appropriate Case Report Forms.

A subject may be withdrawn from the study for the following reasons:

- Subject withdraws consent
- The Investigator requests the subject is withdrawn
- The Medical Monitor or Coordinating Center requests the subject is withdrawn
- The subject’s primary care physician requests the subject is withdrawn
- Non-compliance with the study drug dosing schedule or study visit schedule
- Protocol deviation, protocol violation, or an unanticipated problem
- Lost to follow-up / failure to return for protocol visits
- Early Termination of the Study
- Worsening of another pre-existing disease
- Concurrent (new or worsening) medical condition that interferes with the use of the study drug or completion of the study assessments
  - Clinical significant alteration / abnormality in laboratory values or physical exam after beginning the study (Day 0 and beyond)

### 11.2 Subjects Lost to Follow-up

If a subject is lost to follow-up during the course of the study, the Investigator and / or designated study staff should make reasonable effort to contact the subject and at a minimum encourage the subject to return for an Early Termination Visit to evaluate the subject’s clinical status. All efforts to contact the subject should be documented in the source documents.

## 12. Rules for Termination of the Study

The study may be terminated at any time by the Sponsor. Reasons for terminating the study may include, but are not limited to:

- The incidence or severity of adverse events that indicate a potential health hazard to subjects
- Subject enrollment is unsatisfactory.
- Interim analysis concludes substantial efficacy of the study medication

## 13. Adverse Events

### 13.1 Adverse Event (AE) Definition

An adverse event is any sign, symptom, illness, or medical or psychological condition, not present prior to initiating the study drug, and which develops or worsens during the course of the study, whether or not the event is considered related to the study drug and whether or not the event is expected or unexpected. Medical conditions and diseases present prior to initiating study drug treatment will be considered adverse events only if they worsen after starting treatment with the study drug. Note that Adverse Events related to the known symptoms and clinical course of COVID-19 infection will be tracked separately from those unrelated to COVID-19 infection.

Some examples of adverse events are (but are not limited to):

- A change in the severity, frequency, or duration of a pre-existing condition
- Development of a new, concurrent medical condition
- Development of symptoms, which may or may not be related to the use of a concomitant medication or study drug, or concomitant surgical procedure.
- Laboratory result abnormalities or significant changes, but still within the site reference ranges, following initiation of treatment with the study drug, which the Investigator considers clinically significant.

### 13.2 Serious Adverse Event (SAE) Definition

A serious adverse event is any sign, symptom, illness, or medical or psychological condition, which results in any of the following outcomes:

- Death
- Is life-threatening
- Requires (≥ 24 hours) inpatient hospitalization or prolongs inpatient hospitalization
- Results in persistent or significant disability or incapacity
- Is a congenital anomaly or birth defect
- Is an important medical event - medically significant and which the Investigator regards as serious based on appropriate medical judgment

All Serious Adverse Events, whether thought related or unrelated to study drug, must be reported to the Coordinating Center within 24 hours of the site’s notification of the event; whether or not the Investigator or subject believes the event is related to the study drug. Note that Serious Adverse Events related to the known symptoms and clinical course of COVID-19 infection will be tracked separately from those unrelated to COVID-19 infection. All Serious Adverse Events must be reported to the IRB, in accordance with UVA regulations. If required, follow-up information should also be reported to the Coordinating Center in a timely manner. The Coordinating Center will then prepare the SAE for submission to the appropriate regulatory authorities within the required timeframe.

All Serious Adverse Events which are ongoing at the End-of-Study visit should be followed for an additional 30 days or until resolved, whichever occurs first.

### 13.3 Recording of Adverse Events (AEs) and Serious Adverse Events (SAEs)

All concurrent medical conditions (whether new or worsening) and present prior to or at the Day 0 / Screening Visit (prior to initiating study drug) must be recorded in the Case Report Forms (CRFs) as concurrent medical conditions. All medical conditions, whether new or worsening and present after the initiation of the study drug must be recorded in the CRFs as adverse events (and serious adverse events as indicated), regardless of severity or assessed relationship to the study drug, and reported to the site’s Institutional Review Board (IRB) according to local site IRB guidelines.

At all scheduled and unscheduled study visits, after initiation of the study drug the Investigator or designated personnel should record all voluntary complaints of the subject and directly question the subject regarding the occurrence of any adverse events since their last visit.

All adverse events (serious and not-serious), whether observed by the Investigator or Site Personnel, volunteered by or directly elicited from the subject, and regardless of the relationship with the study drug, should be recorded on the appropriate Case Report Form (the Adverse Event Log). The investigator or designated site personnel, should include a brief description of the adverse event, the date of onset, the date of resolution (when available, or marked as ongoing), the severity of the adverse event, the suspected cause of the adverse event, the possible relationship of the adverse event to the study drug, action taken with the study drug due to the adverse event, and the treatment for the adverse event. The assessment of the event severity and possible relationship to the study drug must be made by the Site Investigator or designee.

### 13.4 Adverse Event (AE) and Serious Adverse Event (SAE) Severity Definitions

The severity of each AE and SAE must be evaluated by the Site Investigator or designee and recorded, as one of the following on the Adverse Event Log:

- **Mild:** Transient (< 48 hours) or mild discomfort; no or minimal medical intervention / therapy required; no or limited interference with the subject’s daily activities.
- **Moderate:** Mild to moderate interference with the subject’s daily activities; possibly none, but usually minimal medical intervention / therapy required.
- **Severe:** Considerable interference with the subject’s daily activities; medical intervention / therapy required; hospitalization possible or likely.

### 13.5 Adverse Event (AE) and Serious Adverse Event (SAE) Relationship to Study Drug Definitions

For each AE and SAE, the relationship to the study drug must be evaluated by the Site Investigator or designee and recorded, as one of the following terms on the Adverse Event Log:

| TERM | DEFINITION | EXPLANATION |
| --- | --- | --- |
| Definite | Clearly related to the study drug | The temporal relationship between the study drug exposure and the onset of the adverse event follows a reasonable sequence; follows a known response pattern to the study drug; and cannot be reasonably explained by the subject's current medical status or by other concomitant therapies / procedures. |
| Probably | Likely related to the study drug | The temporal relationship between the study drug exposure and the onset of the adverse event follows a reasonable sequence; follows a known response pattern to the study drug; and is unlikely to be explained by the subject's current medical status or by other concomitant therapies / procedures. |
| Possibly | May be related to the study drug | The temporal relationship between the study drug exposure and the onset of the adverse event follows a reasonable sequence; follows a known response pattern to the study drug; but may be reasonably explained by the subject's current medical status or by other concomitant therapies / procedures. |
| Unlikely | Likely not related to the study drug | The temporal relationship between the study drug exposure and the onset of the adverse event may follow a reasonable sequence; but the event may be able to be reasonably explained by the subject's current medical status or by other concomitant therapies / procedures. |
| Unrelated | No possible relationship to the study drug | The temporal relationship between the study drug exposure and the onset of the adverse event does not follow a reasonable sequence, and/or the event is most likely explained by the subject's current medical status or by other concomitant therapies / procedures. |

### 13.6 Adverse Event (AE) and Serious Adverse Event (SAE) Analysis

All serious adverse events will be evaluated by the Medical Monitor.

The Data Safety Monitoring Committee (DSMC) will periodically review the blind adverse event and serious adverse event data.

### 13.7 Reportable Events

In addition to Serious Adverse Events, the following other events should be reported to the Coordinating Center within 24 hours of the site’s notification of the event and reported to the site’s Institutional Review Board (IRB) according to local site IRB guidelines:

- Subject withdrawal
- Premature discontinuation of the study drug
- Temporary suspension of the study drug
- Pregnancy
- Major Protocol Violation
- Other events, such as unanticipated problems, which the Site Investigator evaluates as requiring immediate reporting.

## 14. Data and Safety Monitoring Committee (DSMC)

An independent Data Safety Monitoring Committee (DSMC) has been appointed and is responsible for the periodic review and assessment of the study data, with a particular consideration of subject safety through the review of adverse events. The DSMC will meet at least every 2 months (with the first meeting to occur within 2 months of screening of the first subject) throughout the duration of the study, but may convene ad hoc meetings more frequently as required. In reviewing the accumulated study data, the DSMC will determine whether protocol modifications are necessary or if the study should continue without modifications. If the DSMC indicates changes in the protocol should be made, specific recommendations will be made to the Study Principal Investigator and Medical Monitor(s) for consideration and action as required. A report outlining the DSMC’s recommendations will be generated and disseminated after each meeting.

## 15. Statistical Analysis

### 15.1 Primary Objective and Endpoint

The primary study objective is to evaluate the efficacy of daily oral administration of Doxycycline Hyclate 20mg BID as compared with placebo control over the course of 50 days (Day 0 – Day 50) to prevent COVID-19 infection in subjects with ongoing high risk exposure to COVID-19 positive patients.

#### 15.1.1 Power Analysis

Among eligible subjects who have high risk exposure to COVID-19 positive patients, if we randomize subjects in a 1:1 ratio to 50 days of daily oral administration of Doxycycline Hyclate 20mg BID and to 50 days of daily oral administration of placebo, and we have 1938 subjects per study-arm who complete the 50 days of intervention, we expect to have at least 0.80 statistical power to reject the null hypothesis that the risk for COVID-19 infection among subjects who are treated with Doxycycline Hyclate 20mg BID is the same as the risk for COVID-19 infection among subjects who are treated with placebo if the true underlying risk (%) for those who receive Doxycycline Hyclate 20mg BID is less than or equal to 1.25%, and the true underlying risk (%) for those who receive placebo is at least 2.5%. Note that these two stated underlying risks produce an underlying relative risk ratio of 0.5 (Doxycycline Hyclate 20mg BID: Placebo).

#### 15.1.2 Details

The relative risk ratio effect-size of 0.5 was derived by using statistical POWER procedure of SAS version 9.4 (SAS Institute Inc., Cary NC). The input parameters of the POWER procedure were: 1) the underlying reference study population risk of COVID-19 infection; i.e. the probability (p_R_ = 0.025) of COVID-19 infection among subjects who have high risk exposure to COVID-19 positive patients and who are randomized to 50 days of placebo, 2) the underlying non-reference study population risk of COVID-19 infection; i.e. the probability (p_NR_ = 0.0125) of COVID-19 infection among subjects who have high risk exposure to COVID-19 positive patients and who are randomized to 50 days of Doxycycline Hyclate 20mg BID, 3) the power (1-β = 0.80) of the statistical test to reject the null hypothesis that the underlying risk of COVID-19 infection among subjects who have high risk exposure to COVID-19 positive patients is the same for those who receive 50 days of placebo treatment and those who receive 50 days of Doxycycline Hyclate 20mg BID treatment, and 4) the type I error rate (α = 0.05) of the two-sided hypothesis test. Based on these input parameters, the POWER procedure output indicates that we will need a total of 3692 subjects (i.e. 1846 subjects per study-arm).

#### 15.1.3 ower Simulation Result

NA

#### 15.1.4 Subject Withdraw

We anticipate that based on the established safety and tolerability profiles of doxycycline that no more than 5% of the subjects who are determined to be eligible to participate and who are randomized will withdraw from the study. Therefore, we intend to randomize up to 1938 subjects per study-arm (i.e. 3876 in total) in the placebo controlled portion of the study.

### 15.2 Primary Endpoint Analysis

***Primary objective***: The primary objective of this study is to evaluate the efficacy of daily oral administration of Doxycycline Hyclate 20mg BID as compared with placebo control over the course of 50 days to prevent COVID-19 infection in subjects with ongoing high risk exposure to COVID-19 positive patients. ***Primary endpoint:*** The primary endpoint will be the proportion of non COVID-19 infected subjects in each study-arm who go on to become infected with COVID-19 over the course of the 50 day intervention. ***Primary hypothesis:*** The primary null hypothesis will be that the risk for COVID-19 infection among subjects who have high risk exposure to COVID-19 positive patients and who are treated with Doxycycline Hyclate 20mg BID is the same as the risk for COVID-19 infection among subjects who have high risk exposure to COVID-19 positive patients and who are treated with placebo-control: ***Primary analysis:*** In accordance with the stratified randomization scheme of the study (section 5.2.1), the software of the FREQ procedure of SAS version 9.4 (SAS Institute Inc., Cary nC) will be use to conduct a stratified Cochran-Mantel-Haenzel chi-square analysis that will test the null hypothesis that the “common” risk of COVID-19 infection is the same for Doxycycline Hyclate 20mg BID treated subjects and for placebo-control treated subjects (i.e. “common” relative risk ratio = 1), versus the alternative hypothesis that the “common” risk of COVID-19 infection is not the same for Doxycycline Hyclate 20mg BID treated subjects and placebo-control treated subjects (i.e. “common” relative ratio risk ≠ 1). A p≤0.05 decision rule will be used as the null hypothesis rejection criterion for this chi-square test and a 95% confidence interval for the true “common” relative risk ratio will be obtained as part of the FREQ procedure analytical output. ***Test of homogeneity of the risk ratio:*** The “Epi” package of R [7] will be used to conduct a chi-square test to test homogeneity of the relative risk ratio across the different stratification variables (see section 5.2.1). A p≤0.05 decision rule will be used as the null hypothesis rejection criterion for this chi-square test.

### 15.3 Secondary Statistical Analyses

Several secondary endpoints will be analyzed (see Section 2.2.2), and all of the secondary statistical analyses will be conducted in accordance with the guidelines of the intention to treat principle with respect to the randomized clinical study derived data. Open label derived doxycycline cohort data will be included in several secondary analyses and the open label study will be uniquely identified in any secondary analysis in which open label doxycycline cohort data are merged with randomized clinical trial data.

#### 15.3.1 Secondary Endpoint #1 Analysis

***Endpoint:*** Incidence of symptomatic SARS-CoV-2 infections in open label doxycycline, blinded doxycycline, and blinded placebo groups during the Treatment Period. Symptomatic is defined as positive RT-PCR on nasopharyngeal swab with clinical symptoms consistent with presumed COVID-19 infection. ***Primary analysis:*** The Logistic procedure of SAS version 9.4 (SAS Institute Inc., Cary, NC) will be used to conduct an age, sex, race, geographic location, and severity of reported cases in that geographic location at the time of enrollment adjusted analysis of the odds for a positive RT-PCR on nasopharyngeal swab. Adjusted odds will be compared between open label doxycycline treated subjects, blinded doxycycline treated subject, and blinded placebo treated subjects. A p≤0.05 decision rule will be used as the null hypothesis rejection criterion for all between group comparisons. It is important to note that if there are few incidences of symptomatic SARS-CoV-2 infection during the Treatment Period, the “Exact” option of the Logistic procedure will be utilized to estimate the aforementioned adjusted odds ratios using exact methods.

#### 15.3.2 Secondary Endpoint # 2 Analysis

***Endpoint:*** Incidence of asymptomatic SARS-CoV-2 infections in open label doxycycline, blinded doxycycline and blinded placebo group during Treatment Period. Asymptomatic is defined as positive RT-PCR from nasopharyngeal swab without any clinical symptoms associated with COVID-19 infection. ***Primary analysis:*** These data will be analyzed via logistic or exact logistic regression in the same exact way as described in section 15.3.1.

#### 15.3.3 Secondary Endpoint # 3 Analysis

***Endpoint:*** Incidence of symptomatic SARS-CoV-2 infections in open label doxycycline, blinded doxycycline and blinded placebo group in 30 days following Treatment Period. Symptomatic is defined as positive RT-PCR on nasopharyngeal swab with clinical symptoms consistent with presumed COVID-19 infection. ***Primary analysis:*** These data will be analyzed via ordinary or exact logistic regression in the same exact way as described in section 15.3.1.

#### 15.3.4 Secondary Endpoint # 4 Analysis

***Endpoint:*** Incidence of asymptomatic SARS-CoV-2 infections in open label doxycycline, blinded doxycycline and blinded placebo group in 30 days following Treatment Period. Asymptomatic is defined as positive RT-PCR from nasopharyngeal swab without any clinical symptoms associated with COVID-19 infection. ***Primary analysis:*** These data will be analyzed via ordinary or exact logistic regression in the same exact way as described in section 15.3.1.

#### 15.3.5 Secondary Endpoint # 5 Analysis

***Endpoint:*** Evaluation of the incidence of mild to moderate, severe, and critical SARS-CoV-2 infections in open label doxycycline, blinded doxycycline, and blinded placebo group. ***Primary analysis:*** These data will be analyzed via ordinary or exact logistic regression in the same exact way as described in section 15.3.1.

#### 15.3.6 Secondary Endpoint # 6 Analysis

***Endpoint:*** Time-to-first SARS-CoV-2 infection as defined by positive RT-PCR from nasopharyngeal swab. ***Primary analysis:*** Time-to-first SARS-CoV-2 infection will be analyzed by way of Cox proportional hazard regression. *<u>Outcome variable</u>:* Time-to-first SARS-CoV-2 infection will represent the regression outcome variable, with the event times of the subjects who fail to become infected by SARS-CoV-2 right censored at the date of last nasopharyngeal swab test. The predictor variables of the Cox regression model will include a dichotomous variable that will distinguish the subjects who were randomized to doxycycline from the subjects who were randomized to placebo-control, variables to identify the age, sex, and race of the subject, a variable to identify the geographical location of the subject at enrollment, and a variable to identify the severity of reported cases in that geographic location at the time of enrollment. *<u>Hypothesis testing</u>*: The adjusted hazard ratio will serve as the pivotal quantity to compare the adjusted instantaneous risk of SARS-CoV-2 infection between subjects who are randomized to doxycycline and subjects who are randomized to placebo-control. The Wald chi-square test will be used to test the null hypothesis that the adjust hazard ratio is equal to 1; versus the alternative, that the adjusted hazard ratio is not equal to 1. A p≤0.05 decision rule will be used as the null hypothesis rejection criterion for this between group comparison.

#### 15.3.7 Secondary Endpoint # 7 Analysis

***Endpoint***: Time-to-first SARS-CoV-2 clinical event consisting of a persistent change for any of the following: (1) positive nasopharyngeal swab (2) clinical symptoms of COVID-19 infection. ***Primary analysis:*** The time-to-first SARS-CoV-2 clinical event data will be analyzed via Cox proportional hazard regression. The analysis will be conducted in exactly the same way as described in section 15.3.6.

#### 15.3.8 Secondary Endpoint # 8 Analysis

***Endpoint:*** Time-to-first SARS-CoV-2 clinical worsening from asymptomatic to mild to moderate. ***Primary analysis:*** Time-to-first SARS-CoV-2 clinical worsening from asymptomatic to mild to moderate will be analyzed as multiple event time data via the SAS PHREG procedure of SAS version 9.4, with the same model predictor variables as listed in section 15.3.6.

#### 15.3.9 Secondary Endpoint # 9 Analysis

***Endpoint***: **Time-to-first SARS-CoV-2 clinical worsening from mild to moderate to severe.** *Primary analysis:* **Time-to-first SARS-CoV-2 clinical worsening from mild to moderate to severe will be analyzed as multiple event** time data via **the SAS PHREG procedure** of SAS version 9.4, with the same model predictor variables as listed in section 15.3.6.

#### 15.3.10 Secondary Endpoint # 10 Analysis

***Endpoint:*** Rate of subject reported adverse events. *Primary analysis:* Adverse event rates will estimated according to the severity level and compared between the two study arms via Negative binomial regression.

#### 15.3.11 Secondary Endpoint # 11 Analysis

***Endpoint:*** Percentage of subjects who complete the 50 day Treatment Period in all treatment arms. ***Primary analyses:*** These data will simply be summarized by frequencies and percentages.

#### 15.3.12 Secondary Endpoint # 12 Analysis

***Endpoint:*** Percentage subjective compliance of subjects with specified study drug administration schedule. ***Primary analyses:*** These data will simply be summarized by frequencies and percentages.

#### 15.3.13 Secondary Endpoint # 13 Analysis

***Endpoint:*** Percentage objective compliance (as measured by pill counts) of subjects with specified study drug administration schedule. ***Primary analyses:*** These data will simply be summarized by frequencies and percentages.

### 15.4 Safety Data Analysis

Once the first subject in the double blind, placebo controlled study has been randomized, closed and open Data Safety Monitoring reports will be generated at 2 month intervals. Adverse events (AEs) and serious adverse events (SAEs) will be summarized as frequencies and percentages by investigational site, and across investigational sites, for the open report. For the closed report, AEs and SAEs will be summarized according to study arm.

For the open reports, the summaries will be computed and reported for all subjects in the open-label arm. It will then also be reported taking the blinded doxycycline and blinded placebo arms together, without reported the results of these two blinded arms separately. For the closed report the summaries will be and reported according to each separate study arm (i.e. open-label, blinded doxycycline, and blinded placebo each reported separately) without revealing the treatment identity of either of the blinded study arms.

In regard to statistical analyses, if the DSMC requests between study-group comparisons of AE and SAE frequencies, or laboratory results, the comparisons will be provided. Otherwise, the safety data statistical analyses will be conducted via traditional statistical methods following study completion.

### 15.5 Missing Data

Due to the current uncertainty in how to effectively treat COVID-19, and the altruistic mindset of health care workers who will represent the vast majority of the study participants, we anticipate that missing data will be an infrequent event, since the vast majority of the subjects will want to adhere to the study protocol for altruistic reasons and since participate in this study will receive similar tests and examinations as part of their standard of care. However, if a substantial amount of data are missing, the multiple imputation methods of Rubin will be utilized [41]. The MI procedure of SAS version 9.4 (SAS Institute Inc., Cary, NC) will be utilized to impute missing data points, and to also facilitate performing the statistical analyses.

### 15.6 Zinc Sub-analysis

Both primary and secondary endpoint sub-analyses will be performed for those subjects on Zinc supplementation versus those subjects not on Zinc supplementation.

## 16. Regulatory and Ethics Requirements

### 16.1 Statement on Good Clinical Practice (GCP) Compliance

This study will be conducted in accordance with Good Clinical Practices (GCP) using the provisions set forth by the International Conference on Harmonization (ICH) and the United States Food and Drug Administration (FDA), and any applicable national and local regulations. This includes the applicable regulations under 21 Code of Federal Regulations (CFR).

### 16.2 Informed Consent

This study will be conducted in accordance with the provisions set forth by 21 CFR Part 50. The Coordinating Center must be given the opportunity to review the informed consent form prior to submission to the IRB. The Coordinating Center must also be given the opportunity to review any modifications to the informed consent form or informed consent addendums throughout the duration of the study, prior to site submission to the IRB.

Informed consent is a process that is initiated prior to the subject’s agreement to participate in the study and continues through the subject’s study participation. IRB-approved informed consent forms will be provided to the subject and the subject will be asked to read and review the document. The informed consent form must be read and / or explained to each subject by the Investigator or designated site personnel. It is the Investigator’s responsibility to ensure the subject understands the informed consent form, all of the subject’s questions are answered, and that written informed consent form is obtained before the subject participates in any protocol required research procedures. The Investigator is responsible for maintaining the original signed informed consent form and providing the subject with a copy of the informed consent form.

Potential participants will undergo remote HIPAA compliant telemedicine eligibility screening with study staff prior to enrollment. Upon determination of eligibility and signing electronic informed consent documents, participants will be emailed baseline study forms and will be mailed their randomized study treatment. At the end of the study and for their after study visit (Day 50 and Day 80 respectively) participants will received a telemedicine visit to confirm study instructions and that a COViD-19 test has been performed, and be emailed a final study questionnaire. Note that there will be no direct contact between site staff and study subjects in this trial even if local. If there is concern for a potential adverse event necessitating physical exam or other laboratory testing, we will direct the subject to visit their primary care provider or provider responsible for their health care so that they can be evaluated as needed for the purposes of their care and continued participation in the study.

### 16.3 Institutional Review Board (IRB) Review

The Sponsor and Coordinating Center will supply all necessary information and documents to the Investigator for submission of the protocol and informed consent form to the IRB. The Investigator is responsible for providing the appropriate materials to the IRB for review. The study will not commence until the appropriate IRB approval(s) is received. A copy of the approval letter and approved consent form, as well as any other IRB approval documents must be submitted to the Coordinating Center prior to beginning the study.

The Coordinating Center will inform the Investigator of any protocol amendments or required modifications or addendums to the informed consent. The Investigator is responsible for informing the IRB of any amendments to the protocol or informed consent form, and must obtain approval, when required prior to implementing. A copy of the approval letter and approved consent form must be submitted to the Coordinating Center.

The Investigator must ensure all subject recruitment materials are submitted to and approved by the IRB prior to use for subject recruitment. The Investigator is also responsible for reporting all adverse events, protocol deviations and violations, unanticipated problems, and DSMC Reports to the IRB in accordance with IRB guidelines.

The Investigator must obtain a list of the IRB voting members responsible for reviewing the protocol. This list must be updated as necessary throughout the duration of the study. The Investigator is responsible for providing the IRB with study progress reports in accordance with institutional and governmental regulations.

### 16.4 Protocol Amendments

Modifications to the protocol should only be made via a sponsor-issued protocol amendment. Each protocol amendment must be approved by the IRB prior to implementation.

### 16.5 Subject Confidentiality

The Investigator is responsible for maintaining the privacy and confidentiality of subjects at all times, including complying with the obligations to research participants required by the Health Insurance Portability and Accountability Act (HIPAA). To help maintain privacy and confidentiality, subjects will be identified by code numbers on case report forms and other documents submitted to the Medical Monitor and the Coordinating Center and its delegates. However, in the context of the required logistics of the study, given that consent is conducted electronically, email is used directly to the subject for communication, and study drug is sent directly to the subject, the subject will be informed that there is by use of these routes of communication necessarily the potential for exposure of their participation in this study.

However, after the subject signs an informed consent, the site must permit authorized representatives of the Sponsor (this includes the Medical Monitor(s) and Coordinating Center) and federal and local regulatory agencies to review the signed informed consent form and all portions of the subject’s medical record that are directly relevant to the study.

### 16.6 Investigator Responsibilities

The Principal Investigator is responsible for the overall conduct of the study. This includes maintenance of accurate and complete study records, such as source documents, case report forms, IRB records, adverse event records, sponsor correspondence, and regulatory documents. The study Manual of Operations will describe in the detail the responsibilities of the Investigator and will provide a list of study documents that must be maintained.

### 16.7 Monitoring and Regulatory Inspections

This study will be monitored regularly by the Coordinating Center and the Coordinating Center’s authorized delegates. All individuals participating in monitoring activities are in compliance with Good Clinical Practices (GCP). The results of the site monitoring and study progress will be reviewed with the Investigator and other designated site personnel.

### 16.8 Data Management Responsibilities, Source Documents and Case Report Forms (CRFs)

Data collection is the responsibility of the site study personnel, under the supervision of the Principal Investigator. During the study, the Investigator must maintain complete and accurate documentation for the study. The Site must maintain primary source documents supporting all data entered on the Case Report Forms (CRFs). Source documents, include, but are not limited to: documentation of medical and surgical history; study visit record; laboratory results; documentation of internet telemedicine visit or electronic conversations with the subject; demographic information; original, signed informed consent form; records of hospitalization, non-study related procedures, or Emergency Department admissions; documentation of adverse events and changes in medication.

The study will utilize paper Case Report Forms (CRFs). The Sponsor / Coordinating Center will provide the CRFs to the site.

The Coordinating Center will be responsible for data management, quality review, analysis, and reporting of the study data.

### 16.9 Retention of Records

The Investigator is responsible for maintaining intact study records for a period of at least 6 years following the completion of the study. Study records may be retained for a longer period if required by local subject policies. Prior to disposing, changing location, or transferring custody of the study records, the Investigator should contact the Sponsor and/or Coordinating Center.

## List of Abbreviations

AE: Adverse Event
ANCOV: A Analysis of Covariance
BUN: Blood urea nitrogen
CFR: Code of Federal Regulations
CRF: Case Report Form
CRP: C-reactive Protein
DSMC: Data and Safety Monitoring Committee
FDA: Food and Drug Administration
GCP: Good Clinical Practice
HIPAA: Health Insurance Portability and Accountability Act
ICH: International Conference on Harmonization
IRB: Institutional Review Board
MMP: Matrix metalloproteinase
MTX: Methotrexate
RA: Rheumatoid arthritis
RBC: Red Blood Cell
SAE: Serious Adverse Event
SD: Standard Deviation
ULN: Upper Limit of Normal

## Data Availability

The proposed study protocol is provided for informational purposes both with regard to the proposed intervention as well as for the purposes of designing other prospective studies looking at therapeutics for the potential prevention of COVID-19 infection.

## APPENDICES

## Protocol Synopsis

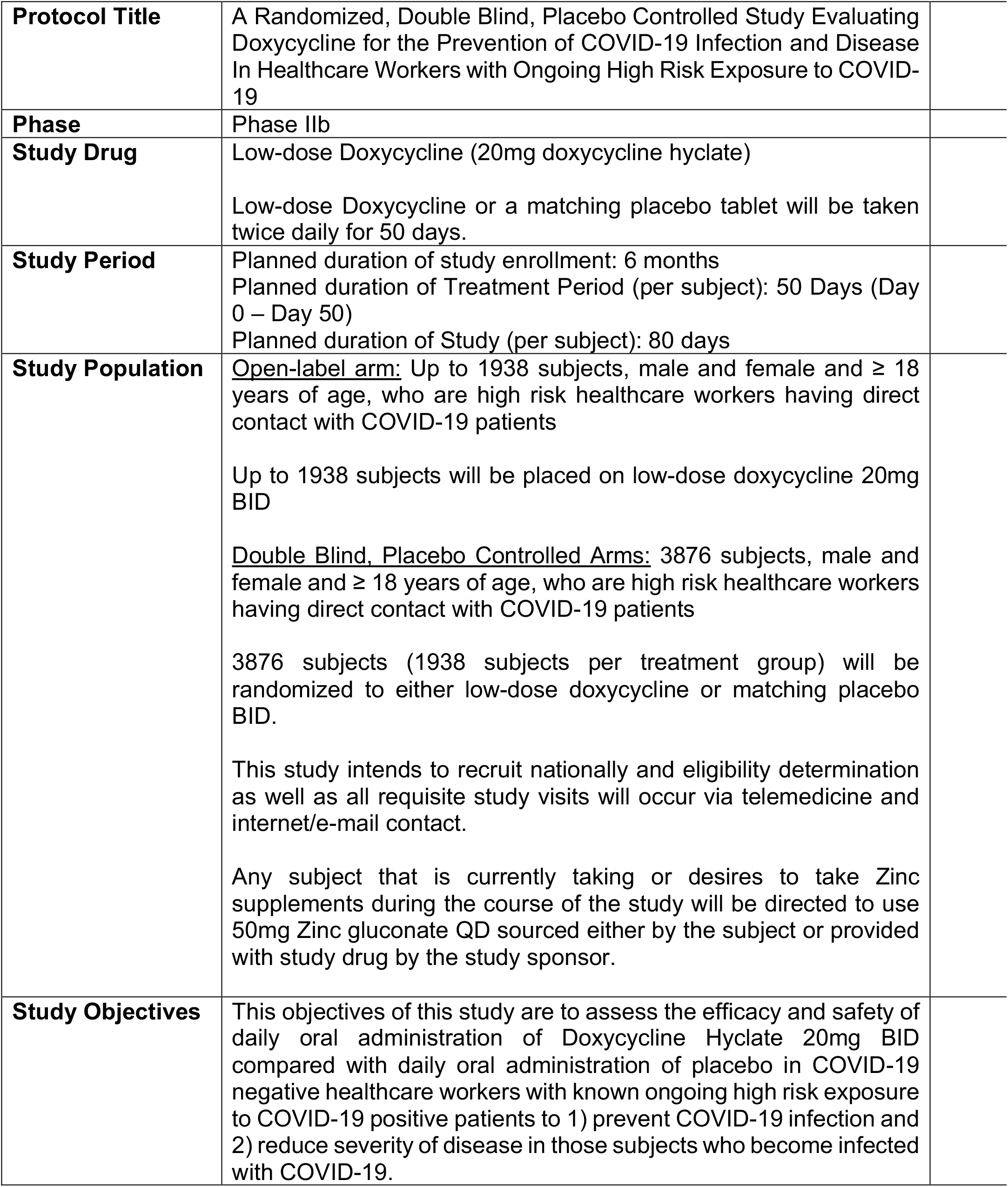

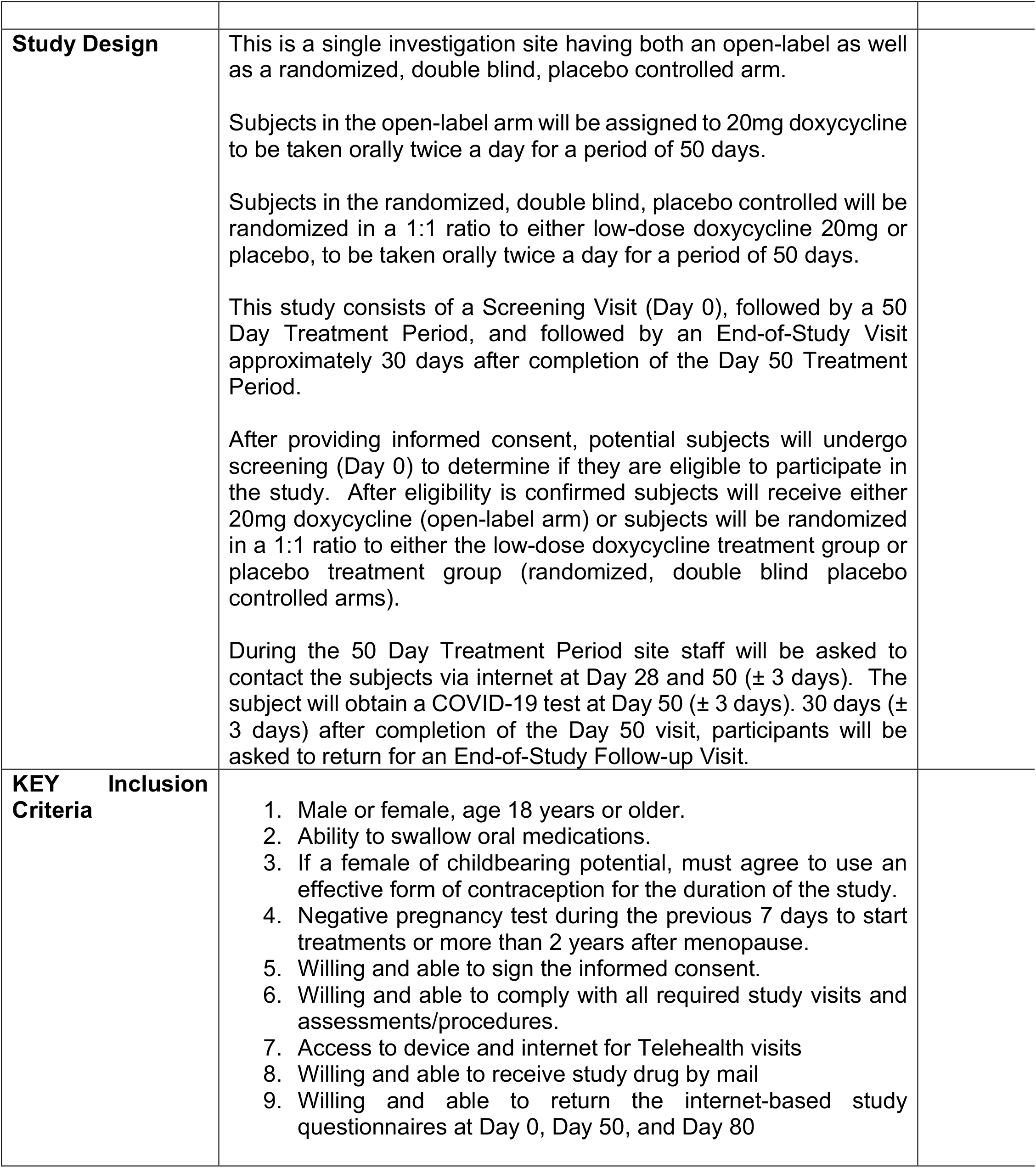

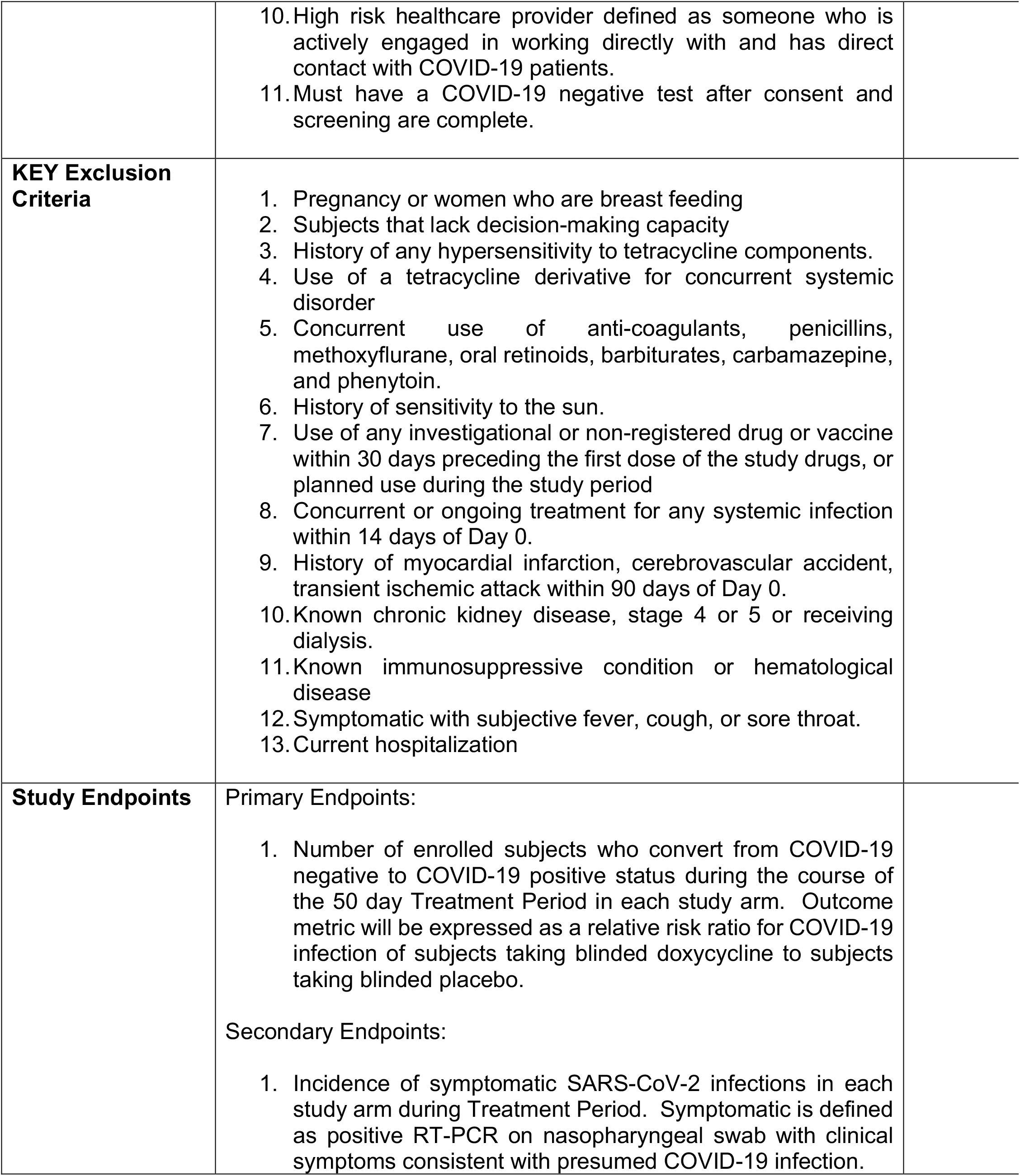

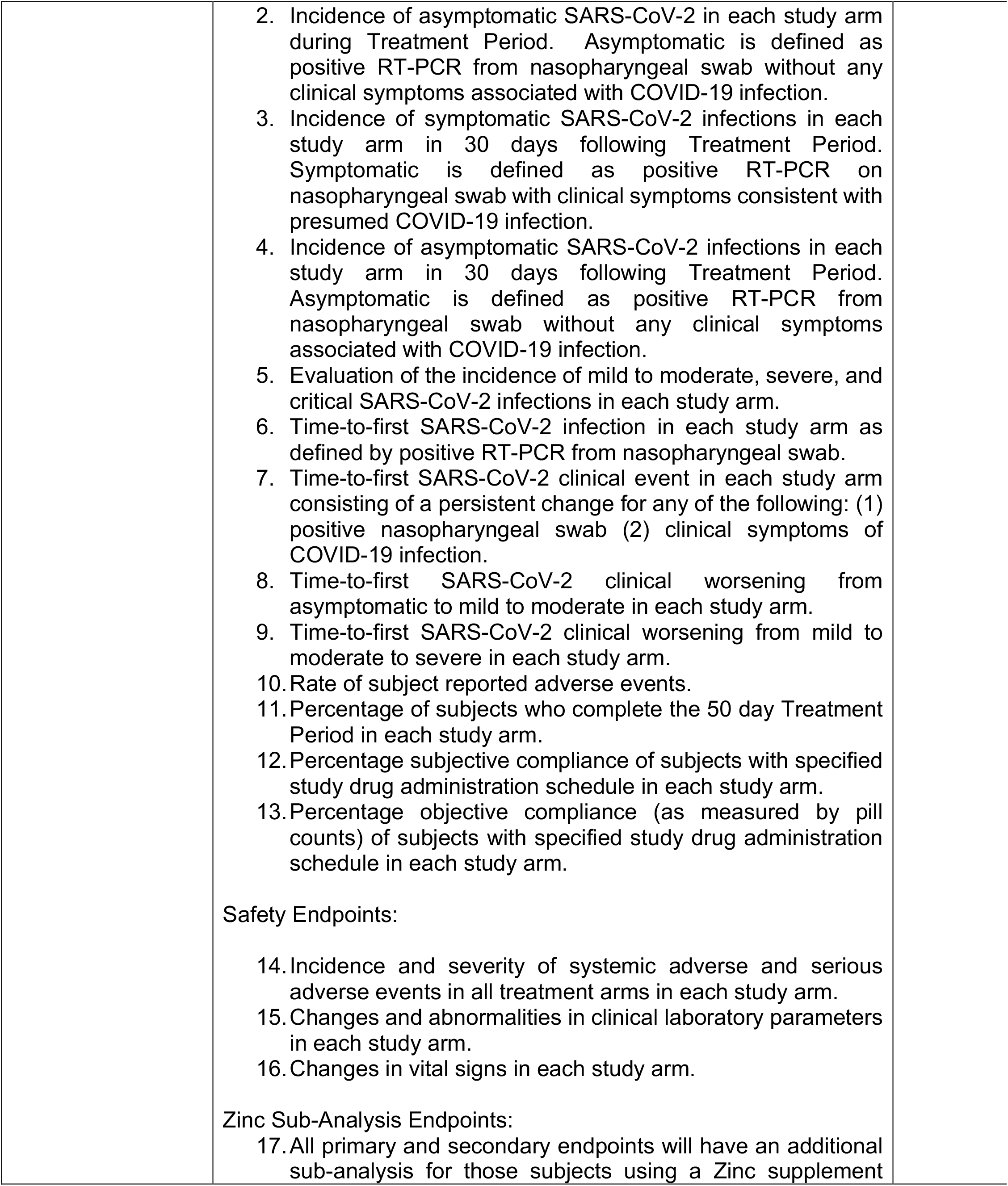

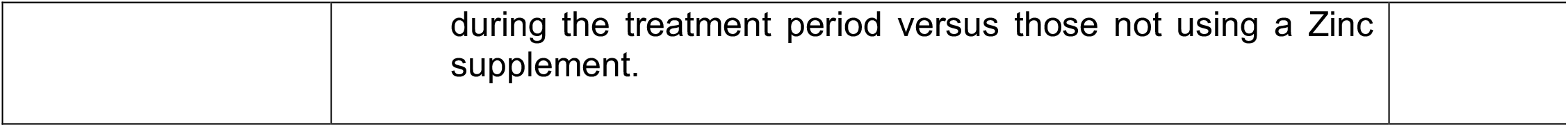

APPENDIX A: Protocol Synopsis
|  |  |
| --- | --- |
| **Protocol Title** | A Randomized, Double Blind, Placebo Controlled Study Evaluating Doxycycline for the Prevention of COVID-19 Infection and Disease In Healthcare Workers with Ongoing High Risk Exposure to COVID-19 |
| **Phase** | Phase IIb |
| **Study Drug** | Low-dose Doxycycline (20mg doxycycline hyclate)<br>Low-dose Doxycycline or a matching placebo tablet will be taken twice daily for 50 days. |
| **Study Period** | Planned duration of study enrollment: 6 months<br>Planned duration of Treatment Period (per subject): 50 Days (Day 0 – Day 50)<br>Planned duration of Study (per subject): 80 days |
| **Study Population** | <u>Open-label arm:</u> Up to 1938 subjects, male and female and $\geq 18$ years of age, who are high risk healthcare workers having direct contact with COVID-19 patients<br>Up to 1938 subjects will be placed on low-dose doxycycline 20mg BID<br><u>Double Blind, Placebo Controlled Arms:</u> 3876 subjects, male and female and $\geq 18$ years of age, who are high risk healthcare workers having direct contact with COVID-19 patients<br>3876 subjects (1938 subjects per treatment group) will be randomized to either low-dose doxycycline or matching placebo BID.<br>This study intends to recruit nationally and eligibility determination as well as all requisite study visits will occur via telemedicine and internet/e-mail contact.<br>Any subject that is currently taking or desires to take Zinc supplements during the course of the study will be directed to use 50mg Zinc gluconate QD sourced either by the subject or provided with study drug by the study sponsor. |
| **Study Objectives** | This objectives of this study are to assess the efficacy and safety of daily oral administration of Doxycycline Hyclate 20mg BID compared with daily oral administration of placebo in COVID-19 negative healthcare workers with known ongoing high risk exposure to COVID-19 positive patients to 1) prevent COVID-19 infection and 2) reduce severity of disease in those subjects who become infected with COVID-19. |
| **Study Design** | This is a single investigation site having both an open-label as well as a randomized, double blind, placebo controlled arm.<br>Subjects in the open-label arm will be assigned to 20mg doxycycline to be taken orally twice a day for a period of 50 days.<br>Subjects in the randomized, double blind, placebo controlled will be randomized in a 1:1 ratio to either low-dose doxycycline 20mg or placebo, to be taken orally twice a day for a period of 50 days.<br>This study consists of a Screening Visit (Day 0), followed by a 50 Day Treatment Period, and followed by an End-of-Study Visit approximately 30 days after completion of the Day 50 Treatment Period.<br>After providing informed consent, potential subjects will undergo screening (Day 0) to determine if they are eligible to participate in the study. After eligibility is confirmed subjects will receive either 20mg doxycycline (open-label arm) or subjects will be randomized in a 1:1 ratio to either the low-dose doxycycline treatment group or placebo treatment group (randomized, double blind placebo controlled arms).<br>During the 50 Day Treatment Period site staff will be asked to contact the subjects via internet at Day 28 and 50 (± 3 days). The subject will obtain a COVID-19 test at Day 50 (± 3 days). 30 days (± 3 days) after completion of the Day 50 visit, participants will be asked to return for an End-of-Study Follow-up Visit. |
| **KEY Inclusion Criteria** | 1. Male or female, age 18 years or older.<br>2. Ability to swallow oral medications.<br>3. If a female of childbearing potential, must agree to use an effective form of contraception for the duration of the study.<br>4. Negative pregnancy test during the previous 7 days to start treatments or more than 2 years after menopause.<br>5. Willing and able to sign the informed consent.<br>6. Willing and able to comply with all required study visits and assessments/procedures.<br>7. Access to device and internet for Telehealth visits<br>8. Willing and able to receive study drug by mail<br>9. Willing and able to return the internet-based study questionnaires at Day 0, Day 50, and Day 80 |
|  | 10. High risk healthcare provider defined as someone who is actively engaged in working directly with and has direct contact with COVID-19 patients.<br>11. Must have a COVID-19 negative test after consent and screening are complete. |
| **KEY Exclusion Criteria** | 1. Pregnancy or women who are breast feeding<br>2. Subjects that lack decision-making capacity<br>3. History of any hypersensitivity to tetracycline components.<br>4. Use of a tetracycline derivative for concurrent systemic disorder<br>5. Concurrent use of anti-coagulants, penicillins, methoxyflurane, oral retinoids, barbiturates, carbamazepine, and phenytoin.<br>6. History of sensitivity to the sun.<br>7. Use of any investigational or non-registered drug or vaccine within 30 days preceding the first dose of the study drugs, or planned use during the study period<br>8. Concurrent or ongoing treatment for any systemic infection within 14 days of Day 0.<br>9. History of myocardial infarction, cerebrovascular accident, transient ischemic attack within 90 days of Day 0.<br>10. Known chronic kidney disease, stage 4 or 5 or receiving dialysis.<br>11. Known immunosuppressive condition or hematological disease<br>12. Symptomatic with subjective fever, cough, or sore throat.<br>13. Current hospitalization |
| **Study Endpoints** | Primary Endpoints:<br>1. Number of enrolled subjects who convert from COVID-19 negative to COVID-19 positive status during the course of the 50 day Treatment Period in each study arm. Outcome metric will be expressed as a relative risk ratio for COVID-19 infection of subjects taking blinded doxycycline to subjects taking blinded placebo.<br>Secondary Endpoints:<br>1. Incidence of symptomatic SARS-CoV-2 infections in each study arm during Treatment Period. Symptomatic is defined as positive RT-PCR on nasopharyngeal swab with clinical symptoms consistent with presumed COVID-19 infection. |
|  | 2. Incidence of asymptomatic SARS-CoV-2 in each study arm during Treatment Period. Asymptomatic is defined as positive RT-PCR from nasopharyngeal swab without any clinical symptoms associated with COVID-19 infection.<br>3. Incidence of symptomatic SARS-CoV-2 infections in each study arm in 30 days following Treatment Period. Symptomatic is defined as positive RT-PCR on nasopharyngeal swab with clinical symptoms consistent with presumed COVID-19 infection.<br>4. Incidence of asymptomatic SARS-CoV-2 infections in each study arm in 30 days following Treatment Period. Asymptomatic is defined as positive RT-PCR from nasopharyngeal swab without any clinical symptoms associated with COVID-19 infection.<br>5. Evaluation of the incidence of mild to moderate, severe, and critical SARS-CoV-2 infections in each study arm.<br>6. Time-to-first SARS-CoV-2 infection in each study arm as defined by positive RT-PCR from nasopharyngeal swab.<br>7. Time-to-first SARS-CoV-2 clinical event in each study arm consisting of a persistent change for any of the following: (1) positive nasopharyngeal swab (2) clinical symptoms of COVID-19 infection.<br>8. Time-to-first SARS-CoV-2 clinical worsening from asymptomatic to mild to moderate in each study arm.<br>9. Time-to-first SARS-CoV-2 clinical worsening from mild to moderate to severe in each study arm.<br>10. Rate of subject reported adverse events.<br>11. Percentage of subjects who complete the 50 day Treatment Period in each study arm.<br>12. Percentage subjective compliance of subjects with specified study drug administration schedule in each study arm.<br>13. Percentage objective compliance (as measured by pill counts) of subjects with specified study drug administration schedule in each study arm.<br>Safety Endpoints:<br>14. Incidence and severity of systemic adverse and serious adverse events in all treatment arms in each study arm.<br>15. Changes and abnormalities in clinical laboratory parameters in each study arm.<br>16. Changes in vital signs in each study arm.<br>Zinc Sub-Analysis Endpoints:<br>17. All primary and secondary endpoints will have an additional sub-analysis for those subjects using a Zinc supplement |
|  | during the treatment period versus those not using a Zinc supplement. |

## APPENDIX B: Study Visit Schedule

|  | Screening / Day 0<br>(± 7 days) | Day 28<br>(± 3 days) | Day 50<br>(± 3 days) | End of Study<br>(30 days ± 7 days) | Early<br>Termination | Un-scheduled Visit |
| --- | --- | --- | --- | --- | --- | --- |
| Informed Consent | X |  |  |  |  |  |
| Demographics | X |  |  |  |  |  |
| Review Study Eligibility<br>(Inclusion / Exclusion Criteria)<br>and Determine Eligibility | X |  |  |  |  |  |
| Medical, Surgical History<br>Review | X |  |  |  |  |  |
| Laboratory Assessments<sup>2</sup> | X |  | X | X | X | X<sup>1</sup> |
| Adverse Event Review |  | X | X | X | X | X |
| Concomitant Medication<br>Review | X | X | X | X | X | X |
| Randomization | X |  |  |  |  | X<sup>1</sup> |
| Study Drug Distribution | X |  | X |  |  | X<sup>1</sup> |
| Study Drug Intake Review |  | X | X | X | X | X<sup>1</sup> |
| Collection of Empty Study Drug<br>Bottles |  |  | X |  | X | X<sup>1</sup> |
| Internet Telemedicine Visit |  | X | X | X | X | X<sup>1</sup> |

1. With the exception of Concomitant Medication Review, Adverse Event Review, and Study Drug Intake Review, all tests and exams to be performed at an Un-scheduled Visit will be determined by the Investigator and are dependent upon the nature of the visit. 2. Laboratory Assessments include: COVID-19 RT-PCR test

